# Rapid review of Allied Health Professionals working in neonatal services

**DOI:** 10.1101/2024.07.23.24310638

**Authors:** Nathan Bromham, Leona Batten, David Jarrom, Elizabeth Gillen, Juliet Hounsome, Jacob Davies, Rhiannon Tudor Edwards, Margaret Manton, Alison Cooper, Adrian Edwards, Ruth Lewis

## Abstract

**Background:** This review aimed to quantify the impact of allied health professionals (AHPs) embedded in neonatal services on outcomes by asking the following review questions:

Q1. What is the effectiveness of neonatal services with embedded allied health professionals compared to neonatal services without embedded allied health professionals?

Q2. What is the effectiveness of early interventions provided by allied health professionals in neonatal units?

**Research Implications and Evidence Gaps:** There was very little directly relevant evidence on AHPs embedded in neonatal services. Most of the evidence related to multidisciplinary team working or early interventions provided by AHPs. Few early intervention trials were from the UK, leading to uncertainty about the availability and applicability of interventions in the UK setting. Further UK-based research is needed to better understand the best way to integrate allied health professionals in neonatal services.

**Economic considerations:** There is no published evidence on the cost of AHPs working within neonatal units. There is marked variability in the reporting of cost estimates for neonatal care units in the UK, making the evaluation of cost implications of adopting AHP recommendations difficult. Subsequent economic evaluations could explore the Budget Impact to the NHS of increasing AHP presence in neonatal units to align with recommendations from AHP professional bodies and Royal Colleges.

**Funding statement:** Health Technology Wales were funded for this work by the Health and Care Research Wales Evidence Centre, itself funded by Health and Care Research Wales on behalf of Welsh Government.

## 1 BACKGROUND

### 1.1 Who is this review for?

This Rapid Review was conducted as part of the Health and Care Research Wales Evidence Centre Work Programme. The review question was suggested by Health Education and Improvement Wales (HEIW). The Maternal Neonatal Safety Support Programme Cymru discovery phase report (PHW 2023) identified that to ensure services are equitable for babies across Wales all neonatal units should have a plan and be allocated capacity to have early developmental intervention using allied health professional (AHP) input. HEIW is undertaking a Strategic Perinatal Workforce Plan programme of work (HEIW 2024) to develop a plan to recruit, retain, train, and transform the perinatal workforce including AHPs. This review is intended to support that work.

### 1.2 Background and purpose of this review

Infants who are critically ill or are born prematurely require medical care in neonatal units to address their immediate physical health needs and ensure survival. These infants also need developmental care, focusing on their longer term neurodevelopmental and psychosocial needs. There can be a perceived trade-off between medical and developmental care for several reasons. Medical care is high intensity with significant staff attention and resources needed – appearing more urgent than developmental care. The busy environment of the neonatal intensive care unit may sometimes interfere with the sleep cycle and sensory development of the infant. Parents and carers may become sidelined if most of their child’s care is provided by healthcare professionals.

Neonatal services, however, increasingly integrate medical and developmental care: an approach which research and guidelines suggest leads to better outcomes for the infant. Examples of this include developmentally supportive care which aims to reduce the stress of the neonatal intensive care unit (NICU) environment on the infant. Family-centred care encourages parents to become involved in the care of their child. Interdisciplinary teams include a range of professionals including allied health professionals who can address all the medical and developmental needs of the infant.

Allied health professionals (AHPs) such as Physiotherapists, Dieticians, Occupational therapists, Speech and language therapists and Psychologists are key members of interdisciplinary/multidisciplinary teams. They provide specialist early interventions and assessments for children in neonatal units. Workforce recommendations for neonatal services (see Table 1) state that AHPs should be embedded as part of the team within neonatal units (BAPM 2022), but currently no Welsh neonatal unit has the recommended whole time equivalent (WTE) number of AHP staff.

**Table 1.**
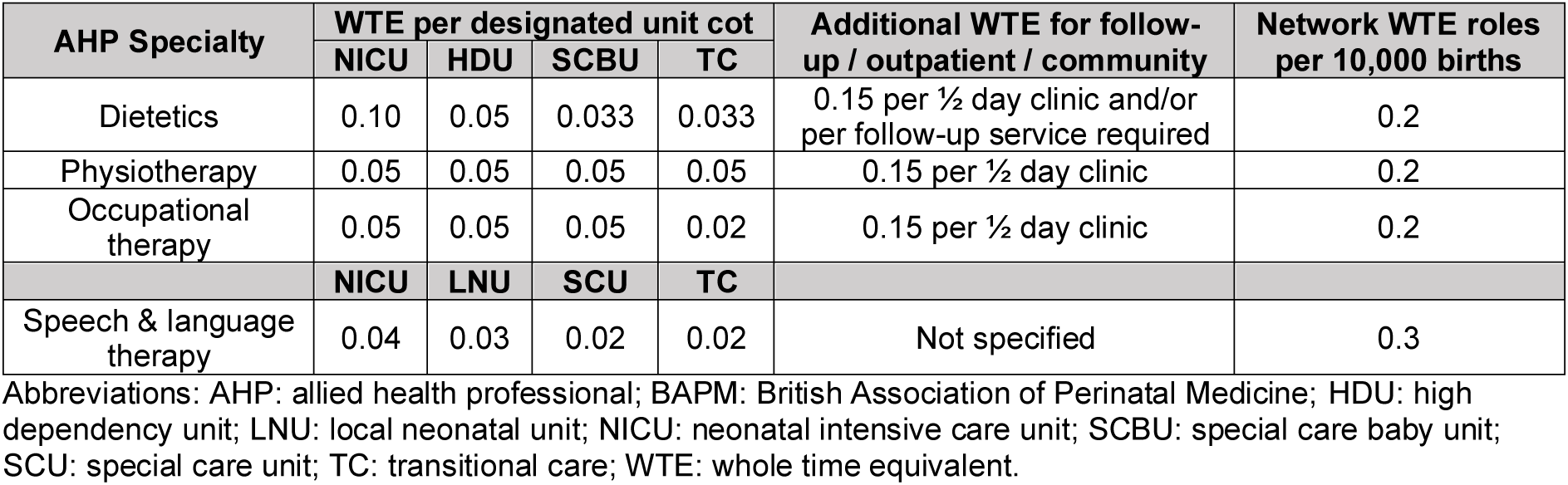
BAPM 2022 AHP staffing recommendations.

This rapid review aims to quantify the impact of the embedded AHP team in neonatal units on outcomes by asking the following review question:

- Q1. What is the effectiveness of neonatal services with embedded allied health professionals compared to neonatal services without embedded allied health professionals?

Preliminary literature searches identified very little evidence comparing AHPs embedded in neonatal services to standard care. For this reason, we asked an additional question, to estimate some of the ways AHPs can impact outcomes in neonatal services:

- Q2. What is the effectiveness of early interventions provided by allied health professionals in neonatal units?

## 2. RESULTS

### 2.1 Q1. Review of neonatal services with embedded allied health professionals

#### Overview of the Evidence Base

Seven studies were included that addressed Q1 (see section 5.1, Table 5 for full eligibility criteria). Six were before-after studies (Furtado et al. 2016, Jeong et al. 2016, Morioka et al. 2024, Muirhead & Bates 2023, Sneve et al. 2008, Welch et al. 2017) and 1 was a comparative cohort study (Gover et al. 2014). The studies were conducted in Canada (n=2), USA (n=2), Australia (n=1). South Korea (n=1) and Japan (n=1). See section 6.2, Table 8 for a detailed summary of the included studies.

Only 1 study (Morioka et al. 2024) directly addressed the review question by comparing before with after the implementation of a neonatal intensive care unit (NICU) with embedded physiotherapy staff. The other studies compared neonatal units with and without multidisciplinary teams including, but not limited to, AHPs. Four of the studies concerned NICU nutritional support multidisciplinary teams with dieticians (Furtado et al. 2016, Gover et al. 2014, Jeong et al. 2016, Sneve et al. 2008). The remaining 2 studies (Muirhead & Bates 2023, Welch et al. 2017) evaluated NICU multidisciplinary teams including occupational therapists, physiotherapists and speech therapists.

All the studies were considered to be at a high risk of including bias in their results or conclusions. This was because of study designs that were more prone to bias, lack of controlling for confounders and lack of published study protocols. (See section 6.3, Table 10 and Table 11 for the summary of the risk of bias assessment for each study).

The certainty of the evidence on which the findings are based has been categorised using the GRADE approach (Grading of Recommendations Assessment, Development and Evaluation) into high, moderate, low or very low. These ratings provide the degree of confidence we have in the findings, with a high rating indicating, that having assessed the potential problems with the available evidence we are very confident that our summary estimate of the intervention effect represents the true value, whilst very low certainly indicates that we have very little confidence that our summary of the effect represents the true underlying effect. Further detail on how studies were assessed is provided in section 6.4.

#### Effectiveness of neonatal services with embedded allied health professionals

**Table 2.** Summary of results and certainty of the evidence for Q1 comparison: neonatal services with embedded AHPs versus services without embedded AHPs.

| Outcomes | No of participants (studies) | Certainty of the evidence (GRADE) | Anticipated absolute effects |  |
| --- | --- | --- | --- | --- |
|  |  |  | Risk with standard care | Risk difference with Neonatal services with embedded AHPs |
| Length of stay assessed with: days | 90 (1 non-randomised study)<sup>a</sup> | Very low<sup>b,c</sup> | The mean length of stay was 121.2 days | MD 0.3 days higher (12.35 lower to 12.95 higher) |
| General gross motor ability assessed with: Hammersmith Neonatal Neurological Examination, total points follow-up: mean 39 weeks PMA | 90 (1 non-randomised study)<sup>a</sup> | Very low<sup>b,c,d</sup> | The mean general gross motor ability was 26.8 points | MD 1.2 points higher (0.15 higher to 2.25 higher) |
| Time to full oral feeding assessed with: weeks PMA | 90 (1 non-randomised study)<sup>a</sup> | Very low<sup>b,c</sup> | The mean time to full oral feeding was 41.0 weeks PMA | MD 1.43 weeks PMA fewer (2.28 fewer to 0.58 fewer) |

Abbreviations. CI: confidence interval; MD: mean difference; PMA: post-menstrual age

**Table 3.** Summary of results and certainty of the evidence for Q1 comparison: neonatal MDTs including allied health professionals versus no such MDT.

| Outcomes | No of participants (studies) | Certainty of the evidence (GRADE) | Relative effect (95% CI) | Anticipated absolute effects |  |
| --- | --- | --- | --- | --- | --- |
|  |  |  |  | Risk with standard care | Risk difference with MDTs with AHPs in neonatal services |
| Mortality | 785 (4 non-randomised studies)<sup>a</sup> | Very low<sup>b,c,d,e,f</sup> | OR ranged from 0.46 to 5.20 | 60 per 1,000 | 60 fewer per 1,000 (31 fewer to 188 more) |
| Length of stay assessed with: days | 785 (4 non-randomised studies)<sup>a</sup> | Very low<sup>b,c,d,f</sup> | - | The median length of stay was 73.4 days | MD 10 days lower (0 to 0 ) |
| Parental confidence assessed with: Parents contribute to care planning | 97 (1 non-randomised study)<sup>g</sup> | Very low<sup>b,d,e</sup> | RR 1.61 (1.09 to 2.40) | 413 per 1,000 | 252 more per 1,000 (37 more to 578 more) |
| Time to oral feeding assessed with: days | 722 (3 non-randomised studies)<sup>h</sup> | Very low<sup>b,c,d,f</sup> | - | The median time to oral feeding was 34 days | 0 days (0 to 0 ) |

Abbreviations. CI: confidence interval; MD: mean difference; OR: odds ratio; RR: risk ratio

##### Length of stay

Very low certainty evidence from 1 before-after study (Morioka et al. 2024) suggests that NICU with embedded physiotherapists compared to standard physiotherapy care does not affect length of stay in NICU.

##### General gross motor ability

Very low certainty evidence from 1 before-after study shows (Morioka et al. 2024) a benefit in terms of Hammersmith Neonatal Neurological Examination total points score for infants cared for in a NICU with embedded physiotherapists compared to standard physiotherapy care.

##### Oral feeding

Very low certainty evidence from 1 before-after study (Morioka et al. 2024) that infants cared for in a NICU with embedded physiotherapists reached full oral feeding around 1½ weeks sooner (post menstrual age) than infants receiving standard physiotherapy care.

##### Mortality

Very low certainty evidence from 3 before-after studies (Furtado et al. 2016, Jeong et al. 2016, Sneve et al. 2008) and 1 cohort study (Gover et al. 2014) shows inconsistent results in terms of mortality when comparing infants cared for by a multidisciplinary nutritional support team including a dietician with those not cared for by such a team. Study results ranged from clinically important benefit to clinically important harm with the nutritional support MDT and it was not appropriate to pool the results.

##### Length of stay

Very low certainty evidence from 3 before-after studies (Furtado et al. 2016, Jeong et al. 2016, Sneve et al. 2008) and 1 cohort study (Gover et al. 2014) shows inconsistent results in terms of length of stay when comparing infants cared for by a multidisciplinary nutritional support team including a dietician with those not cared for by such a team. Study results ranged from clinically important benefit to clinically important harm with the nutritional support MDT and it was not appropriate to pool the results.

##### Parental confidence

Very low certainty evidence from 1 before-after study (Muirhead & Bates 2023) showed that NICU staff rated parents as much more likely to contribute to care planning following implementation of multidisciplinary developmental care rounds including an advanced practice nurse, an occupational therapist, a physiotherapist, and a speech therapist.

##### Oral feeding

Very low certainty evidence from 2 before-after studies (Furtado et al. 2016, Jeong et al. 2016) and 1 cohort study (Gover et al. 2014) shows inconsistent results in terms of time to full oral feeding when comparing infants cared for by a multidisciplinary nutritional support team including a dietician with those not cared for by such a team. Study results ranged from clinically important benefit to clinically important harm with the nutritional support MDT and it was not appropriate to pool the results.

#### Bottom line results for review of neonatal services with embedded allied health professionals

Evidence from a single before-after study suggests AHPs embedded in neonatal services may improve gross motor ability and lead to earlier oral feeding, but this evidence is uncertain. Results from before-after and cohort studies of multidisciplinary nutrition support teams in neonatal units are too inconsistent to draw conclusions. There was a lack of evidence on general cognitive ability, quality of life, visual or hearing impairment, parental bonding, attachment, mental health or mood outcomes.

### 2.2 Q2. Review of early interventions provided by allied health professionals in neonatal units

#### Overview of the evidence base

Five studies were included that addressed Q2 (see section 5.1, Table 6 for full eligibility criteria) all were systematic reviews (Girabent-Farrés et al. 2021, Greene et al. 2023, Lavallée et al. 2021, Orton et al. 2024, Rodovanski et al. 2023). The systematic reviews were checked for trials relevant to our review (where the early intervention was delivered by an AHP while the infant was in a neonatal unit) and 57 relevant randomised controlled trials (RCTs) were identified. See Table 9 for a summary of the included systematic reviews and relevant trials.

The systematic reviews focused on different types of early intervention including: early interventions actively involving parents (Girabent-Farrés et al. 2021, Lavallée et al. 2021), oral stimulation (Greene et al. 2023), early interventions which continue post-discharge (Orton et al. 2024) and multisensory stimulation (Rodovanski et al. 2023).

The systematic reviews were at low (Greene et al. 2023, Lavallée et al. 2021, Orton et al. 2024) or unclear (Girabent-Farrés et al. 2021, Rodovanski et al. 2023) risk of bias (see Table 12). Reasons for unclear risk of bias were concerns with the search strategy and inability to access the review protocol. This means that some relevant trials may have been missed by the systematic reviews.

The risk of bias judgements for the individual trials were also extracted from the systematic reviews and used to inform certainty in the body of evidence (see Forest plots in Appendix 1).

The main risk of bias issue in the randomised trials was lack of blinding, although some of the oral or multisensory stimulation trials did have blinding. Many of the trials were at high or unclear risk of bias from allocation concealment.

#### Effectiveness of early interventions provided by AHPs in neonatal services

The effectiveness evidence comes from RCTs identified in the included systematic reviews that were relevant to our review question.

**Table 4.** Summary of results and certainty of the evidence for Q2 comparison: AHP delivered or supported early intervention versus standard care.

| Outcomes | No of participants (studies) | Certainty of the evidence (GRADE) | Anticipated absolute effects |  |
| --- | --- | --- | --- | --- |
|  |  |  | Risk with standard care | Risk difference with AHP delivered or supported early intervention |
| Length of hospital stay (oral stimulation studies) assessed with: days | 534 (10 RCTs)<sup>a</sup> | Moderate<sup>b</sup> | The median length of hospital stay (oral stimulation studies) was 45 days | MD 6.61 days lower (8.52 lower to 4.71 lower) |
| Length of hospital stay (TKS studies) assessed with: days | 570 (13 RCTs)<sup>c</sup> | Low<sup>b,d</sup> | The median length of hospital stay (TKS studies) was 25.5 days | MD 1.73 days lower (3.22 lower to 0.25 lower) |
| Length of hospital stay (ATVV studies) assessed with: days | 923 (9 RCTs)<sup>c</sup> | Very low<sup>b,d,e</sup> | The median length of hospital stay (ATVV studies) was 30.1 days | MD 0.06 days lower (1.85 lower to 1.73 higher) |
| Parental bonding and attachment assessed with: Parental Sensitivity | 247 (3 RCTs)<sup>f</sup> | Moderate<sup>g,h</sup> | - | SMD 0.32 SD higher (0.07 higher to 0.57 higher) |
| General cognitive ability assessed with: developmental quotient | 743 (7 RCTs)<sup>i</sup> | Moderate<sup>b</sup> | - | SMD 0.22 SD higher (0.07 higher to 0.36 higher) |
|  |  |  | Risk with standard care | Risk difference with AHP delivered or supported early intervention |
| General gross motor ability assessed with: developmental quotient | 786 (8 RCTs)<sup>i</sup> | Moderate<sup>b</sup> | - | SMD 0.32 SD higher (0.18 higher to 0.46 higher) |
| Time to oral feeding (oral stimulation studies) assessed with: days | 409 (10 RCTs)<sup>a</sup> | Low<sup>b,j</sup> | The median time to oral feeding (oral stimulation studies) was 20 days | MD 7.63 days lower (10.5 lower to 4.75 lower) |
| Parental stress (short term) assessed with: PSI or PSI-SF | 179 (2 RCTs)<sup>k</sup> | Low<sup>b,l</sup> | - | SMD 0.07 SD lower (0.37 lower to 0.22 higher) |

Abbreviations. CI: confidence interval; MD: mean difference; SMD: standardised mean difference

#### Length of stay

Moderate certainty evidence from 10 RCTs (Greene et al. 2023) indicates that length of stay is around 1 week shorter with oral stimulation provided by an AHP compared to standard care.

Low certainty evidence from 13 RCTs (Rodovanski et al. 2023) indicates that length of stay is around 2 days shorter with tactile-kinaesthetic stimulation compared to standard care.

Very low certainty evidence from 9 RCTs (Rodovanski et al. 2023) suggests no difference between length of stay with auditory-tactile-visual-vestibular intervention and with standard care.

#### Parental bonding and attachment

Moderate certainty evidence from 3 RCTs (Lavallée et al. 2021) shows a small beneficial effect of early interventions delivered by psychologists for parents of preterm babies during NICU stay compared to standard care, in terms of parental sensitivity.

#### General cognitive ability

Moderate certainty evidence from 7 RCTs (Orton et al. 2024) shows a small beneficial effect of early interventions delivered by AHPs compared to standard care, in terms of developmental quotient in infancy. The estimated improvement is around 3 points on the developmental quotient scale, where 100 is the score for a typically developing infant.

#### General gross motor ability

Moderate certainty evidence from 8 RCTs (Orton et al. 2024) shows a small beneficial effect of early interventions delivered by AHPs compared to standard care, in terms of developmental quotient in infancy. The estimated improvement is around 5 points on the developmental quotient scale, where 100 is the score for a typically developing infant.

#### Oral feeding

Moderate certainty evidence from 10 RCTs (Greene et al. 2023) indicates that time to oral feeding is around 1 week shorter with oral stimulation provided by an AHP than with standard care.

#### Parental stress

Low certainty evidence from 2 RCTs (Girabent-Farrés et al. 2021) found no important difference in short-term parental stress when comparing early interventions actively involving parents delivered by AHPs to standard care.

#### Bottom line results for review of early interventions provided by allied health professionals in neonatal units

Moderate certainty evidence from RCTs suggests that early interventions provided by AHPs in neonatal units are associated with shorter length of stay, better parental sensitivity and quicker oral feeding. These interventions are also associated with small improvements in general cognitive and general gross motor ability in infancy compared to standard care.

There is low certainty evidence from RCTs that early interventions delivered by AHPs do not impact parental stress in the short-term. There was a lack of evidence on aspiration, cranial head shape, readmission rates, neonatal staff competencies and neonatal staff stress outcomes.

## 3. DISCUSSION

### 3.1 Summary of the findings

The aim of this rapid review was to summarise the effectiveness of neonatal services with embedded AHPs compared to neonatal services without embedded AHPs. While minimal directly relevant evidence was found—most of it concerned multidisciplinary team working— the available evidence did support the inclusion of embedded AHPs

The second review question provided indirect evidence to support the place of AHPs in neonatal services with 57 RCTs identified that largely supported the effectiveness of early interventions delivered by AHPs in neonatal units.

### 3.2 Strengths and limitations of the available evidence

Most of the studies of neonatal services with embedded AHPs were before-after designs. These are at high risk of biases such as trends over time, confounders and regression to the mean. This means that differences in outcomes between the before and after periods may not solely be due to the intervention.

The before-after studies of nutritional support MDTs with dieticians showed a great deal of heterogeneity in their results. This may be because they included somewhat different populations such as infants with gastroschisis, short bowel syndrome or with very low birthweight. In addition, these interventions were primarily aimed at nutritional outcomes such as weight gain. While these studies typically showed benefits for these nutritional outcomes these were not outcomes of interest in this review and so were not included in the evidence.

In the studies of neonatal units with and without MDTs, AHPs were part of the team with a range of other healthcare professionals so it is not possible to assess their individual impact on outcomes.

The majority of the included studies were conducted in neonatal intensive care units (level 3 neonatal units) and it is unclear how it applies to local level 1 or 2 neonatal units were infants may have less complex needs.

The included RCTs of early interventions provided by AHPs covered all types of AHPs in our review protocol (occupational therapists, physiotherapists, speech and language therapists and psychologists) – except for dieticians. This may be because trials of early interventions provided by dieticians did not specifically mention the involvement of the dietitian in the systematic review publication.

The early intervention evidence on cognitive and motor outcomes comes from RCTs that started in the neonatal unit, but the interventions carried on post-discharge. It is unclear how this might apply to the responsibilities of AHPs embedded in neonatal units and to what extent they are involved in the care of the infant post-discharge.

In the evidence on multisensory early interventions carried out in NICU it was difficult to identify exactly who carried out the intervention due to poor reporting. The evidence was downgraded for indirectness, as the interventions were likely delivered by nurses in some cases.

The focus of this review was the involvement of AHPs in services and early interventions rather than which specific early interventions are the most effective. For this reason different types of intervention have been pooled within general classes of intervention. This is likely to contribute to heterogeneity in some cases. A range of interventions were reported in the evidence but it is uncertain how many of these interventions are available in the UK setting. Three included trials were from the UK.

We had anticipated that relevant evidence would come from UK audits of neonatal services. While a national neonatal audit programme (RCPCH 2023) was identified it did not provide data on AHPs and no other relevant audits were found. We also looked for evidence about staffing numbers or AHP to cot ratios to inform the optimal number of AHPs in a neonatal unit, but no relevant evidence was found.

### 3.3 Strengths and limitations of this Rapid Review

One of the strengths of this review is the systematic and comprehensive search of the literature. This identified a dearth of evidence on the effectiveness of neonatal services with embedded allied health professionals. This is despite national policy and guidance which recognises the importance of AHPs in neonatal services and states AHPs should be embedded in these services.

This review likely underestimates the role of the AHP in neonatal services. Many of the interventions provided in neonatal units are provided using a team approach which becomes part of routine care, such as environmental light or sound modifications, massage, skin to skin care, use of maternal voice or positioning. While the AHP may support these approaches their role is not typically documented as a specific service in trials of these interventions so they would have been excluded from the evidence. Similarly, the Newborn Individualized Developmental Care and Assessment Program (NIDCAP) model of care is a team-based approach that is part of routine care and would be supported by AHPs – but NIDCAP trials typically describe the intervention as being delivered by nurses.

The evidence shows benefits of early interventions delivered by AHPs in neonatal services and if a large part of the AHP’s time is spent in the neonatal unit it may be practical to embed them there. The benefit of AHPs in neonatal services can then extend beyond delivering early interventions themselves. For instance, their presence, advice and training can positively impact other neonatal staff and parents, contributing to a culture of developmental care. AHPs working in neonatal units are typically required to be neonatal specialists (APCP Neonatal Committee 2023, British Dietetic Association 2022, Royal College of Occupational Therapists 2022, Royal College of Speech and Language Therapists 2023), embedding AHPs within units enables them to reach these specialist levels.

### 3.4 Implications for policy and practice

The GIRFT neonatology workforce national report (GIRFT 2022) recommends “Embed allied health professionals, pharmacy and psychology services into neonatal units and networks, in line with professional standards to improve outcomes for babies.” Clinical guidelines from NICE (NICE 2017, NICE 2020) indicate that AHPs are an essential part of care and follow-up for children born preterm that are being cared for in neonatal units.

National neonatal service and quality standards from BAPM (BAPM 2022) set out the staffing service standards for AHPs in neonatal units. UK AHP professional bodies and Royal Colleges (APCP Neonatal Committee 2023, Association of Clinical Psychologists 2022, British Dietetic Association 2022, Royal College of Occupational Therapists 2022, Royal College of Speech and Language Therapists 2023) have each made specific recommendations about the number of WTE AHP staff of each type needed for each level of neonatal unit (largely based on expert consensus). Findings of this rapid review support the idea that the involvement of AHPs neonatal units is likely to improve outcomes, but it does not inform the exact numbers of staff required.

Implementing the recommendations of BAPM, the AHP professional bodies and Royal Colleges in Welsh neonatal units is likely to be a major change in practice as no Welsh neonatal unit currently has the recommended WTE number of AHP staff. This may be related to a challenging climate in neonatal staffing – the National Neonatal Audit Programme (NNAP) 2022 audit (RCPCH 2023) found only 71% of neonatal nurse shifts were staffed according to recommended levels in England, Wales and Scotland. The GIRFT neonatology workforce national report (GIRFT 2022) found significant shortfall in AHP service provision in England with less than half of neonatal services having regular dietetics, physiotherapy, speech and language therapy and occupational therapy services. In units with AHP services, adherence to staffing standards was very low – the highest being dietetics at only 54% adherence.

It is recognised that additional funding and planning are needed to meet the challenge of AHP staffing in neonatal services (NHSE & NHSI 2019). The 2019 NHS Long Term Plan committed to additional funding (up to 2024) to meet the action plan of the NHS England Neonatal Critical Care Review (NCCR). One of the Long Term Plan’s key commitments was “Further developing the expert neonatal workforce required: extra neonatal nurses and expanded roles for some allied health professionals to support clinical care.” To implement the AHP recommendations of the NCCR, NHS England (NHSE & NHSI 2019) advised “NHS Trusts should develop an AHP strategy as part of workforce planning which sets out the level and expertise of pharmacy and AHP required, the level currently available, and how any gaps will be filled.”

In 2022 the Welsh Government asked Improvement Cymru undertake the Discovery Phase for a new Maternity and Neonatal Safety Support Programme (PHW 2023). This project identified adherence to national workforce standards as a key priority. It acknowledged that national workforce planning needs a strategy to ensure that AHPs are embedded within services in line with national standards. It also highlighted the role of AHP leads within the Maternity and Neonatal Network. HEIW (HEIW 2024) is developing a strategic perinatal workforce plan to recruit, retain, train and transform the current and future perinatal workforce in NHS Wales.

### 3.5 Implications for future research

Further research is needed on the effective organisation of AHPs within UK neonatal services, including staffing levels, expanding roles and service models.

### 3.6 Economic considerations*

**Cost of neonatal units**

- There is marked variability in the reporting of cost estimates for neonatal care units in the UK. With reporting differing due to study heterogeneity, differences in data sources, costing methodologies and care requirements for babies (Yang et al. 2023).

**Directions for future research**

- Future research should assess the costs of introducing AHPs in neonatal units. Evaluations should comprehensively consider the costs of AHP staff of each type and band, as defined in recommendations from AHP professional bodies and Royal Colleges. The latest unit costs for AHPs in the UK are presented in the Unit Costs of Health and Social Care 2023 (Jones et al. 2023).
- Future research should focus on the relative cost-effectiveness of neonatal units that embed AHPs within their operating model and those that do not. Consideration should be given to the impact of neonatal care not just on babies but upon the wider family including the mother.
- Subsequent economic evaluations could explore the Budget Impact to the NHS of increasing AHP presence in neonatal units to align with recommendations from AHP professional bodies and Royal Colleges.

## Data Availability

All data produced in the present study are available upon reasonable request to the authors

## Abbreviations

Acronym: Full Description
AHP: Allied health professional
BAPM: British Association of Perinatal Medicine
ATVV: Auditory-Tactile-Visual-Vestibular
CBT: Cognitive Behavioural Therapy
CI: Confidence Interval
EMDR: Eye Movement Desensitisation and Reprocessing
EPOC: Effective Practice and Organisation of Care
GRADE: Grading of Recommendations, Assessment, Development and Evaluation
HDU: High Dependency Unit
ICN: Intensive Care Nurse
MD: Mean Difference
MDT: Multidisciplinary Team
NHSE: NHS England
NHSI: NHS Improvement
NICU: Neonatal intensive care unit
OR: Odds Ratio
PMA: Post Menstrual Age
PSI: Parenting Stress Index
PSI-SF: Parenting Stress Index – Short Form
RCT: Randomized Controlled Trial
ROB: Risk of Bias
SCBU: Special Care Baby Unit
SMD: Standardized Mean Difference
TC: Transitional Care
TDPCI: Therapist-Delivered Postural Control Intervention
TKS: Tactile-Kinaesthetic Stimulation
WTE: Whole Time Equivalent

## 5. RAPID REVIEW METHODS

### 5.1 Eligibility criteria

**Table 5:** Eligibility Criteria for review question 1: what is the effectiveness of neonatal services with embedded allied health professionals compared to neonatal services without embedded allied health professionals?

|  | **Inclusion criteria** |
| --- | --- |
| **Population** | Babies admitted to neonatal units and their parents/carers |
| **Intervention** | Neonatal units with the following embedded AHPs:<br>• Physiotherapists<br>• Dieticians<br>• Occupational therapists<br>• Speech and language therapists<br>• Psychologists |
| **Control or counter intervention** | • Neonatal units without embedded AHPs<br>• Neonatal units with different ratios of embedded AHPs to babies cared for<br>• Neonatal units with different types of embedded AHPs |
| **Outcome measures** | Primary outcomes:<br>• Mortality<br>• General gross motor ability<br>• General cognitive ability<br>Secondary outcomes:<br>• Quality of life<br>• Length of stay<br>• Visual impairment<br>• Hearing impairment<br>• Parental confidence<br>• Parental bonding and attachment<br>• Parental mental health and mood |
| **Setting / Context** | Neonatal units (levels 1 to 3) |
| **Study design** | Systematic reviews of comparative studies<br>Randomised controlled trials<br>Comparative observational studies |
| **Countries** | We will prioritise papers that include data from countries where evidence is most relevant to UK neonatal services. We will classify this as studies from UK, Europe, Australia, New Zealand and Canada. Evidence from other settings, may be included but will be downgraded for indirectness. |
| **Language of publication** | English |
| **Publication date** | 1995 onwards |
| **Publication type** | Published and preprint. Audits and other grey literature may be included if they are publicly available. |

**Table 6:** Eligibility Criteria for review question 2: What is the effectiveness of early interventions provided by allied health professionals in neonatal units?

|  | Inclusion criteria | Exclusion criteria |
| --- | --- | --- |
| **Population** | Babies admitted to neonatal units and their parents/carers |  |
| **Intervention** | Early interventions provided in neonatal units by AHPs such as:<br>• Physiotherapist<br>○ Handling<br>○ Head moulding/shaping<br>○ Neonatal massage<br>○ Early Therapy in Perinatal Stroke (eTIPS)<br>• Dietician<br>○ Introduction of enteral feeds<br>○ Optimisation of nutrition<br>○ Introduction of nutrition protocols<br>○ Introduction of breast milk fortifier<br>• Speech and language therapist<br>○ feeding / pre-feeding strategies<br>▪ cue-based feeding/infant-driven feeding<br>▪ pacing<br>▪ slow flow teat<br>▪ elevated side-lying<br>○ neonatal communication focused interventions<br>• Occupational therapist<br>○ Infant occupations<br>○ Parent occupations<br>○ Parent/infant co-occupations<br>• Psychologist<br>○ Emotional / cognitive support interventions:<br>▪ Emotional grounding<br>▪ Normalising<br>▪ Active listening<br>▪ Emotional stabilisation<br>▪ Mindfulness<br>▪ Sharing formulations<br>▪ Early sensitivity training<br>○ Specific therapeutic interventions:<br>▪ Eye Movement Desensitisation and Reprocessing (EMDR)<br>▪ Cognitive Behavioural Therapy (CBT)<br>○ Attachment and Relationships based interventions<br>• Interventions not exclusive to a single type of AHP:<br>○ Developmentally supportive care<br>○ Positioning<br>○ Education (of parents and neonatal staff)<br>○ Coaching of parents<br>○ Sensory stimulation<br>○ Non-pharmacological pain relief<br>○ Specialist assessments | Interventions where the involvement of an AHP is unclear |
| **Control or counter intervention** | • Standard care<br>• No early intervention |  |
| **Outcome measures** | Primary outcomes:<br>• Length of stay<br>• Parental confidence<br>• Parental bonding and attachment<br>• General gross motor ability<br>• General cognitive ability<br>Secondary outcomes (some are profession specific):<br>• Aspiration<br>• Oral feeding<br>• Cranial head shape<br>• Readmission rates<br>• Parental mental health and mood<br>• Neonatal staff competencies<br>• Neonatal staff stress |  |
| **Setting / Context** | Neonatal units (levels 1 to 3) |  |
| **Study design** | Systematic reviews of randomised controlled trials<br>Randomised controlled trials |  |
| **Countries** | We will prioritise papers that include data from countries where evidence is most relevant to UK neonatal services. We will classify this as studies from UK, Europe, Australia, New Zealand and Canada. Evidence from other settings, may be included but will be downgraded for indirectness. |  |
| **Language of publication** | English |  |
| **Publication date** | 1995 onwards |  |
| **Publication type** | Published and preprint |  |

### 5.2 Incorporating data from existing reviews

For review question 2 we used included systematic reviews:

As a source of study characteristics/patient demographics. As a source of outcome and risk of bias data.

### 5.3 Literature search

Medline, Ovid EMCARE, AMED, CINAHL and Cochrane were searched for review question 1. For review question 2 Ovid EMCARE, AMED, CINAHL and Cochrane were searched. Studies published after 1995 were included due to major advances in neonatal care since that date. Searches were done between 7^th^ and 11^th^ March 2024. See Appendix 2 for search strategy. Websites were also searched for audits of neonatal services, see Appendix 3.

### 5.4 Study selection process

Using the Covidence systematic reviewing platform, two reviewers dual-screened 10% of titles and abstracts independently. After this, the level of agreement was assessed with disagreements settled by discussion and consensus. Both reviewers had to achieve at least 80% agreement on screened records before progressing to the next stage. The remaining titles and abstracts were screened by the primary reviewer alone. Ten percent of all full texts were screened by both reviewers, with the same agreement threshold (80%) as before, which was necessary before the remaining records could be screened by the primary reviewer alone. During independent screening, the primary reviewer consulted with the secondary reviewer in the case of any uncertainties.

### 5.5 Data extraction

The following data were extracted where available:

Study information (author, year, country, type of neonatal unit) Population characteristics

Intervention and comparison characteristics

Data on relative effectiveness for example odds ratios, or risk ratios for dichotomous variables and mean differences for continuous variables.

Data were extracted by a single reviewer and quality assured by a second reviewer.

### 5.6 Study design classification

The included studies were classified as systematic reviews, RCTs, cohort studies or before-after studies.

### 5.7 Quality appraisal

Different critical appraisal checklists were used to assess risk of bias depending on study design. For systematic reviews we used ROBIS (Whiting et al. 2016). For observational cohort studies the ROBINS-I checklist (Sterne et al. 2016) was used. ROBINS-I was not appropriate for before-after studies so in these cases we used the EPOC risk of bias criteria (EPOC 2017). We used the included systematic reviews as a source of risk of bias data for trials in review question 2. These reviews had used Cochrane RoB 1, Cochrane RoB 2 and the PEDro scale to assess risk of bias in RCTs trials.

### 5.8 Synthesis

Studies were grouped by intervention and outcome for synthesis. When studies were sufficiently similar in their populations, interventions, comparisons and outcomes we combined their results in meta-analysis using the meta package in R (Balduzzi et al. 2019). Otherwise, the data were synthesised using narrative synthesis. Published meta-analyses of the included systematic reviews were redone, excluding any studies not meeting our inclusion criteria.

### 5.9 Assessment of body of evidence

Certainty in the overall body of evidence was assessed using GRADE (Guyatt et al. 2008). The GRADE approach provides a formal system to categorises the certainty of (or confidence in) the evidence for each outcome or effect into one of four levels: high, moderate, low and very low. The system for assessing the certainty of evidence includes five domains relating to: risk of bias across the studies, inconsistency (heterogeneity) of the results between studies, indirectness (including subgroup analyses and applicability of the outcome measure), imprecision of the result (number of events, and width of the confidence intervals), and publication bias. At the start it is assumed that randomised controlled trials (RCTs) provide high level evidence and observational studies low, but this evidence is then downgraded or upgraded based on the assessment of the different criteria. A summary of the GRADE ratings and their interpretation is provided in Table 7.

**Table 7.** GRADE ratings and their interpretation.

| Grade (evidence quality) | Definition |
| --- | --- |
| **High** | We are very confident that the true effect lies close to that of the estimate of the effect |
| **Moderate** | We are moderately confident in the effect estimate: The true effect is likely to be close to the estimate of the effect, but there is a possibility that it is substantially different |
| **Low** | Our confidence in the effect estimate is limited: The true effect may be substantially different from the estimate of the effect. |
| **Very low** | We have very little confidence in the effect estimate: The true effect is likely to be substantially different from the estimate of effect |

## 6. EVIDENCE

### 6.1 Search results and study selection

**Figure 1:**
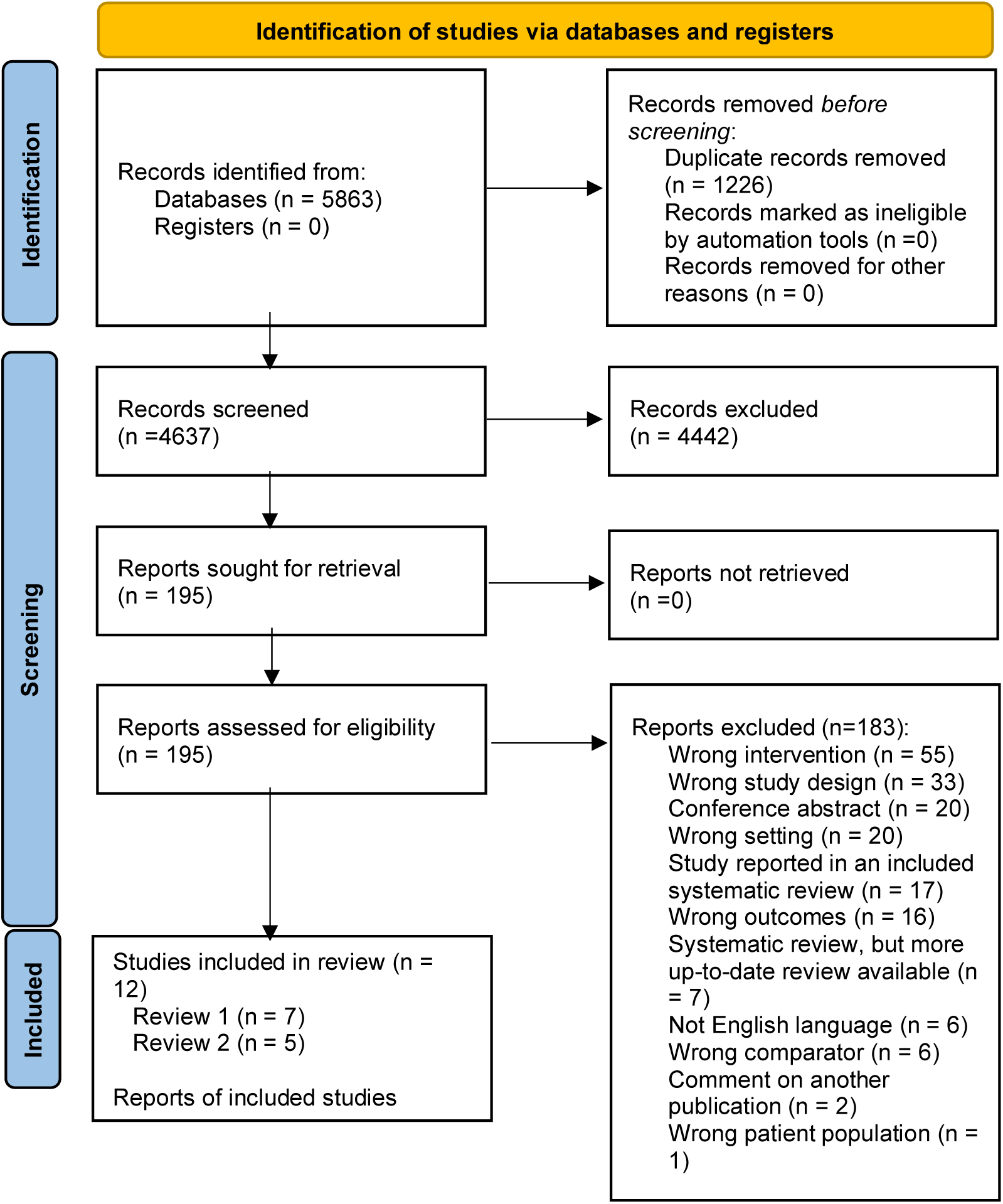
PRISMA flow diagram.

### 6.2 Data extraction

Studies included for question 1 are summarised in Table 8, all were primary studies. Studies included for question 2 are summarised in Table 9, all were systematic reviews.

**Table 8:**
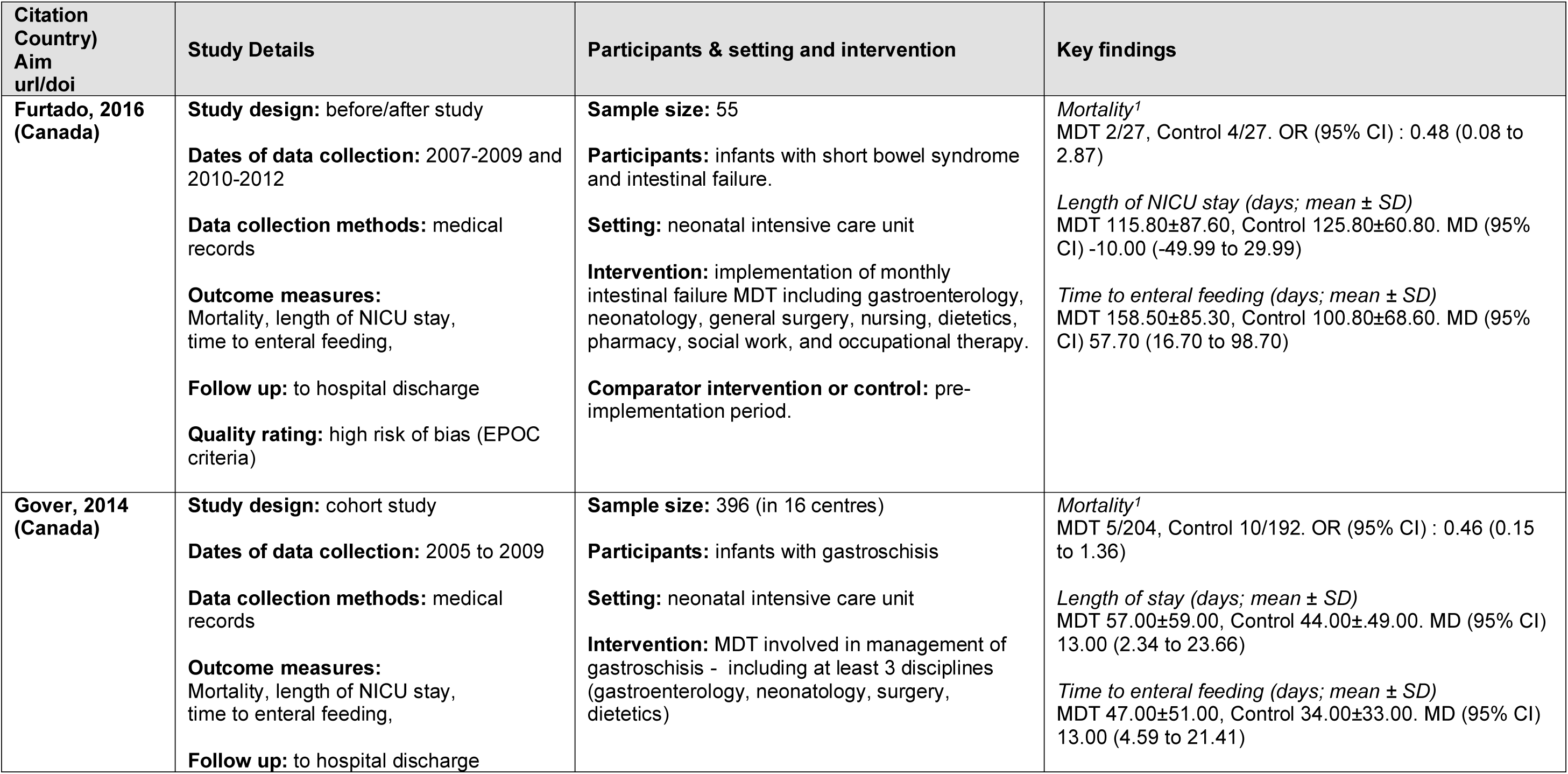
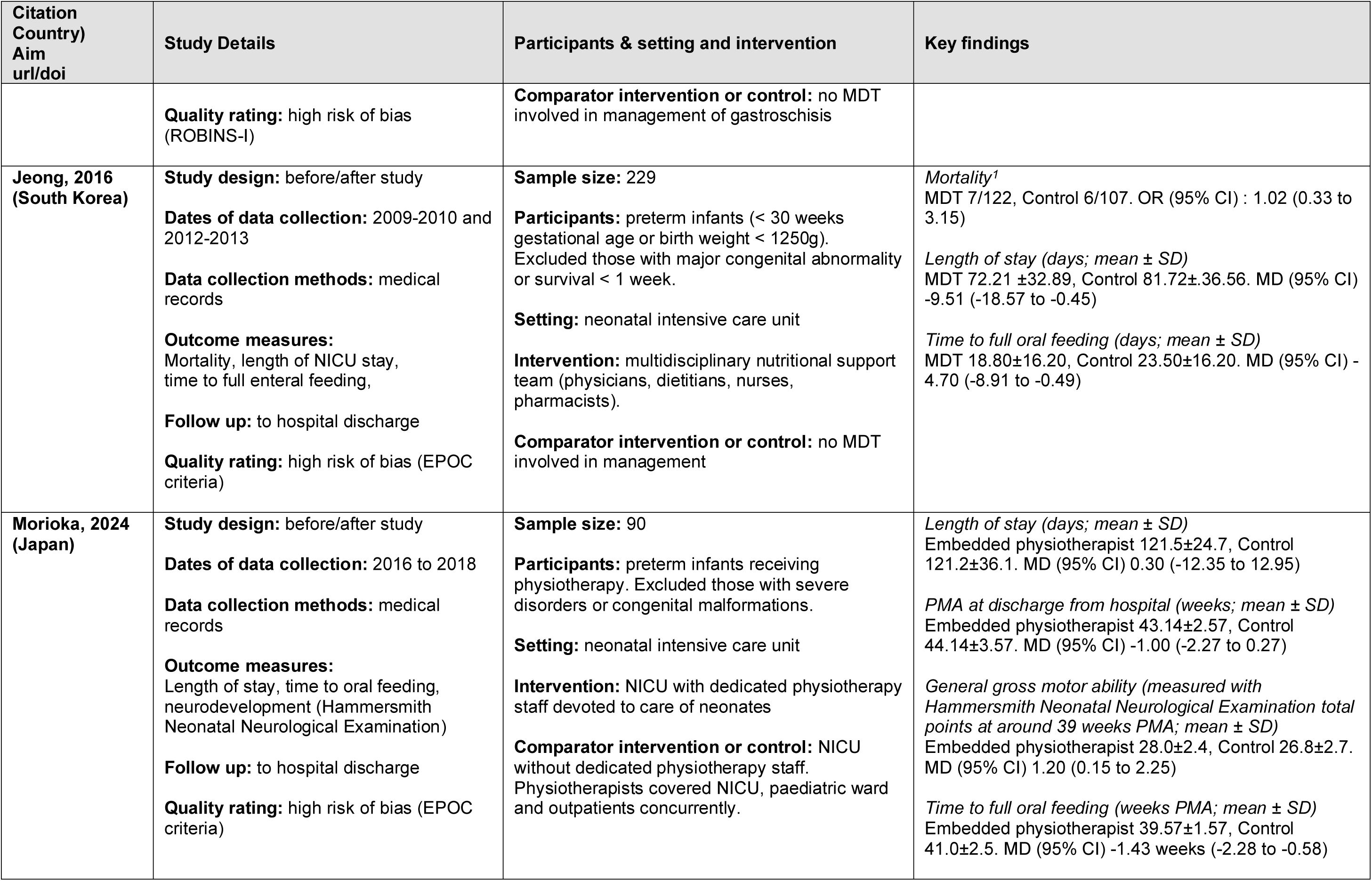
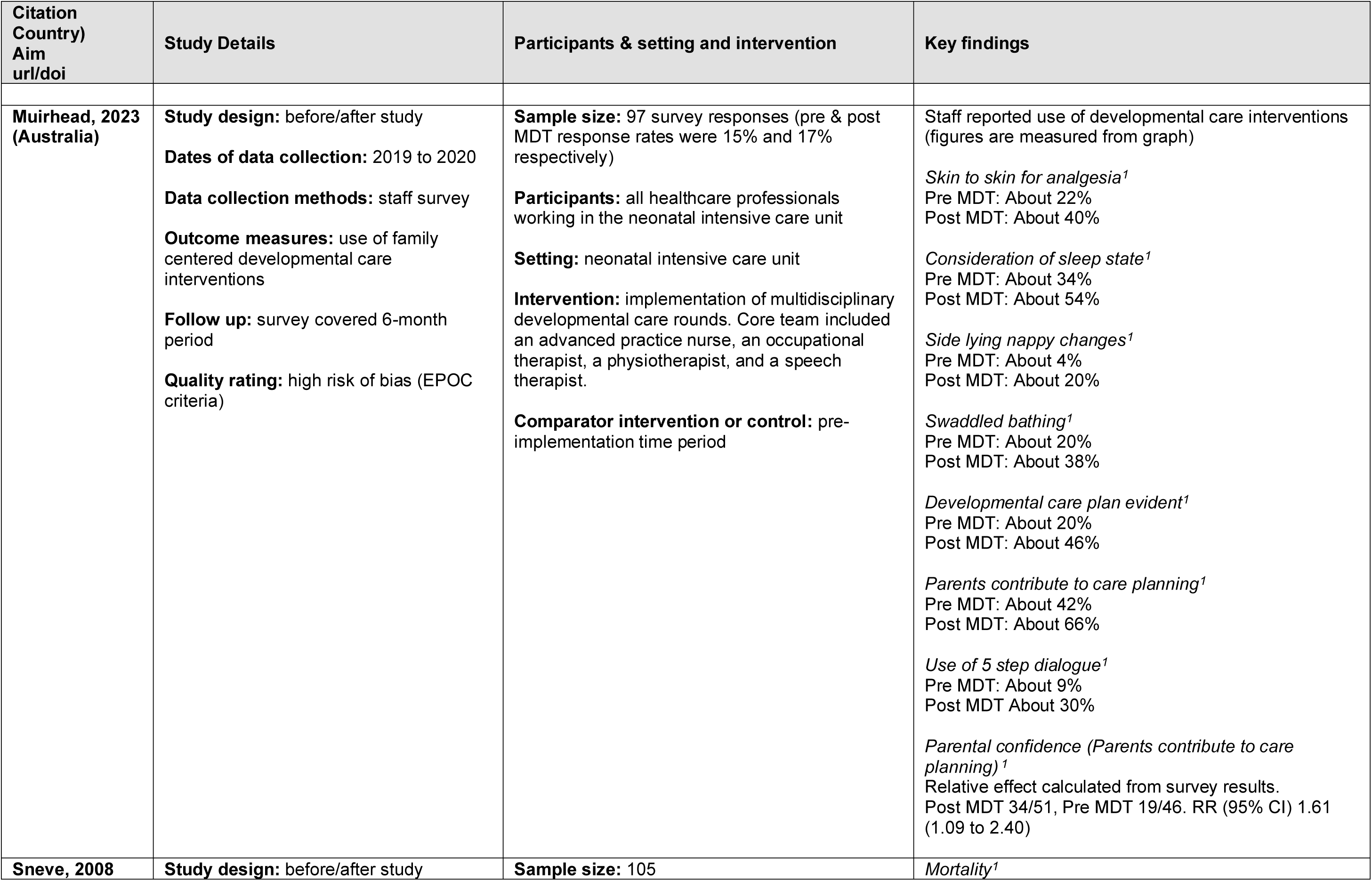
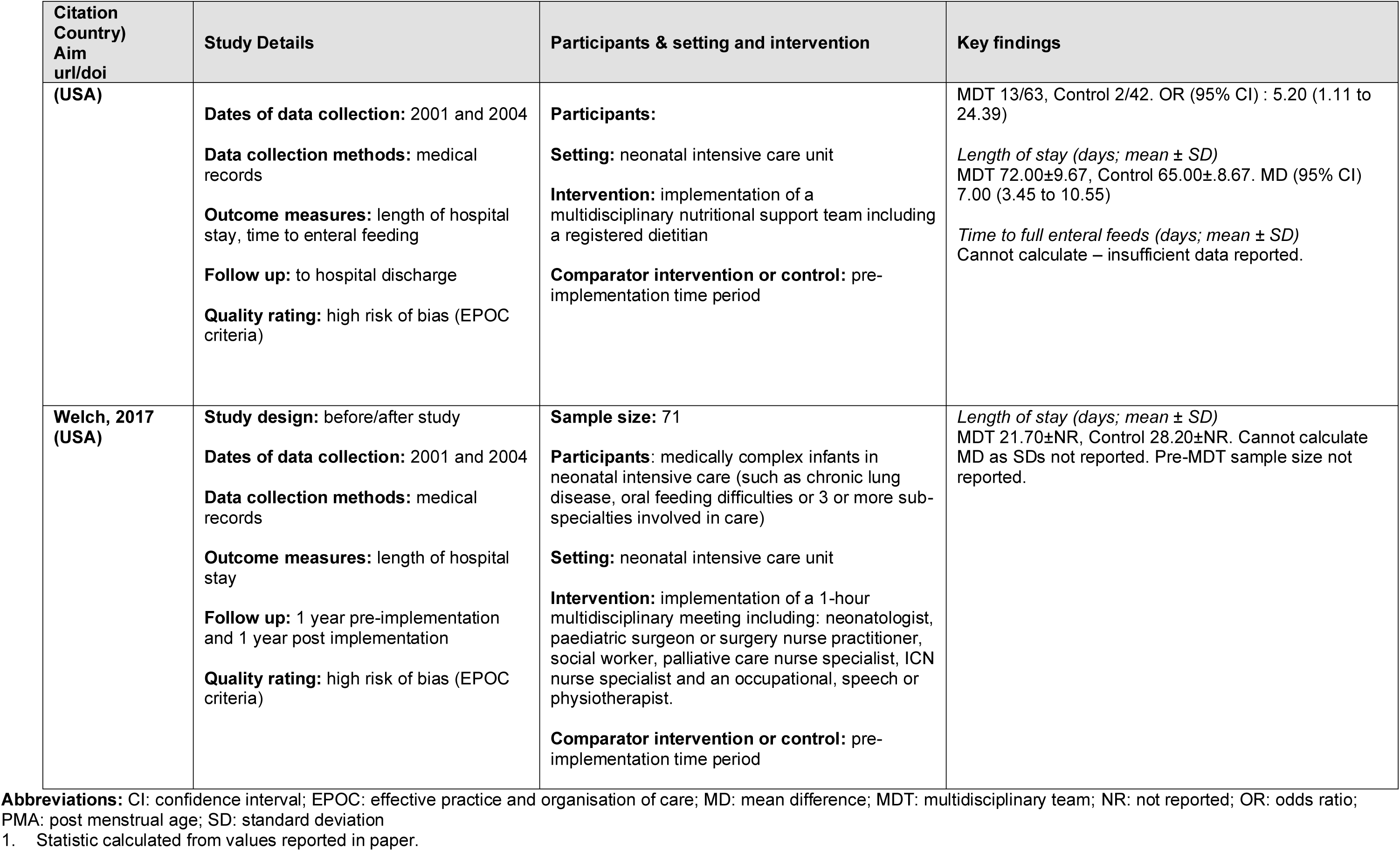
Summary of included studies for Q1.

**Table 9:**
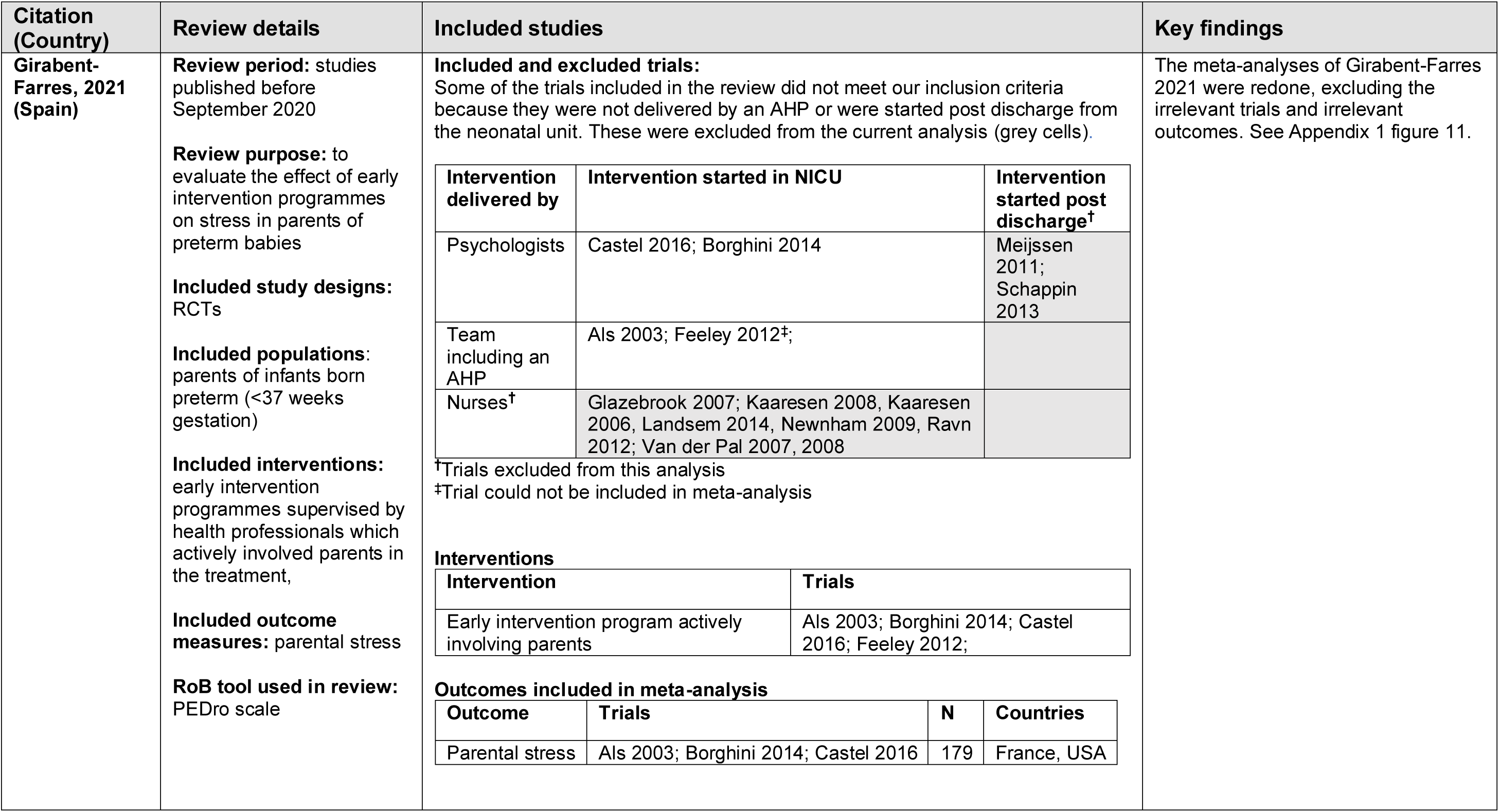
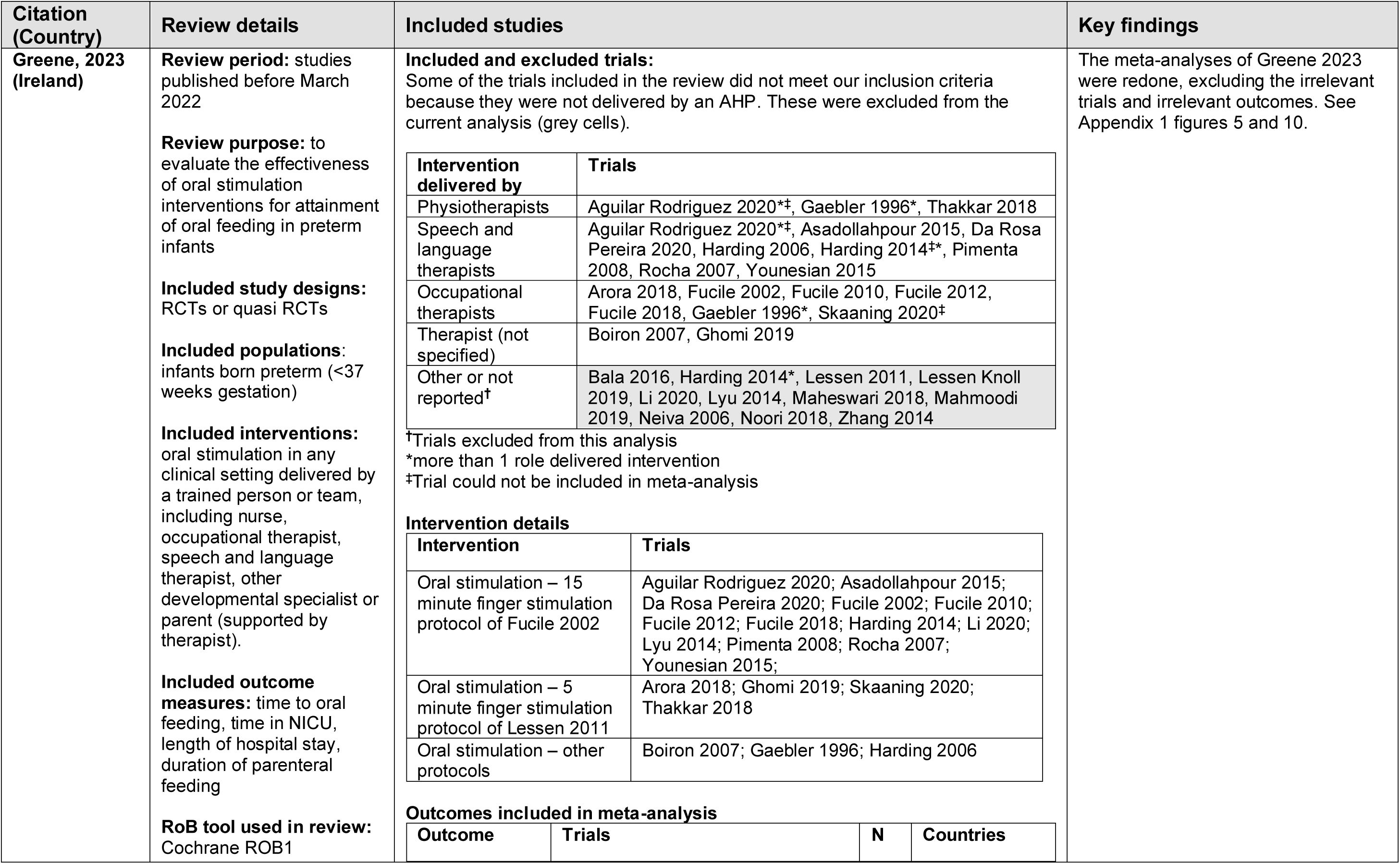
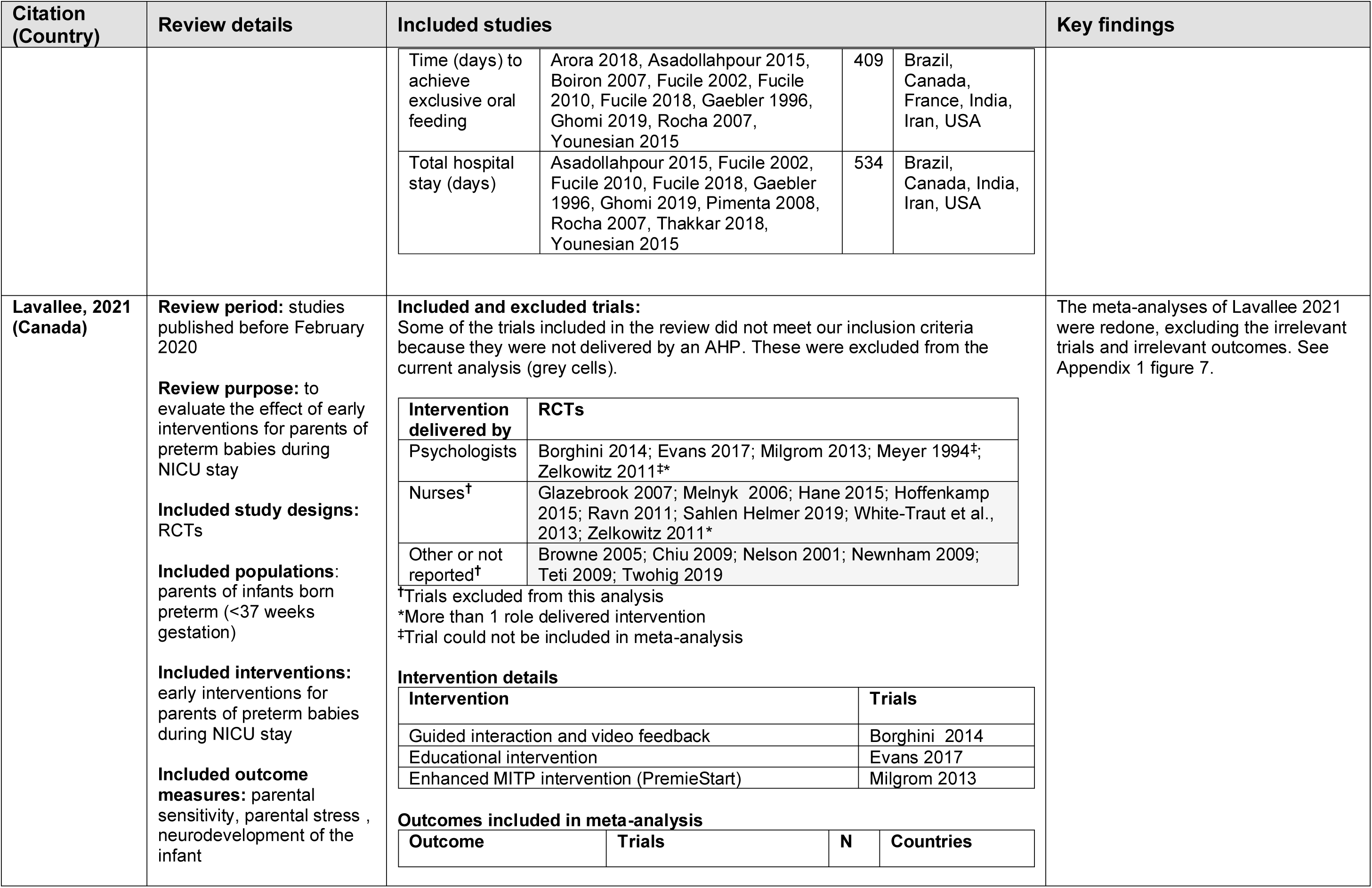
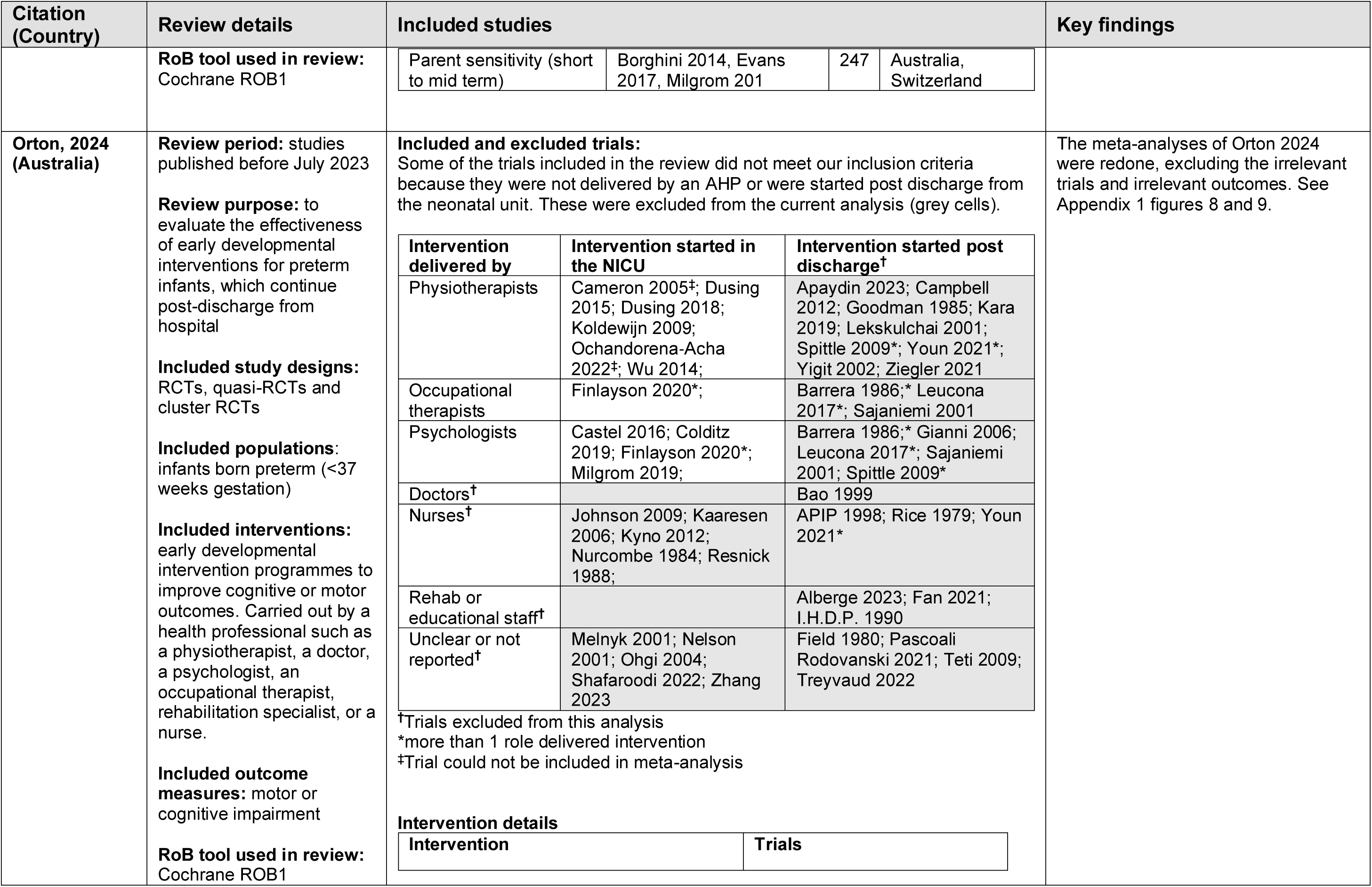
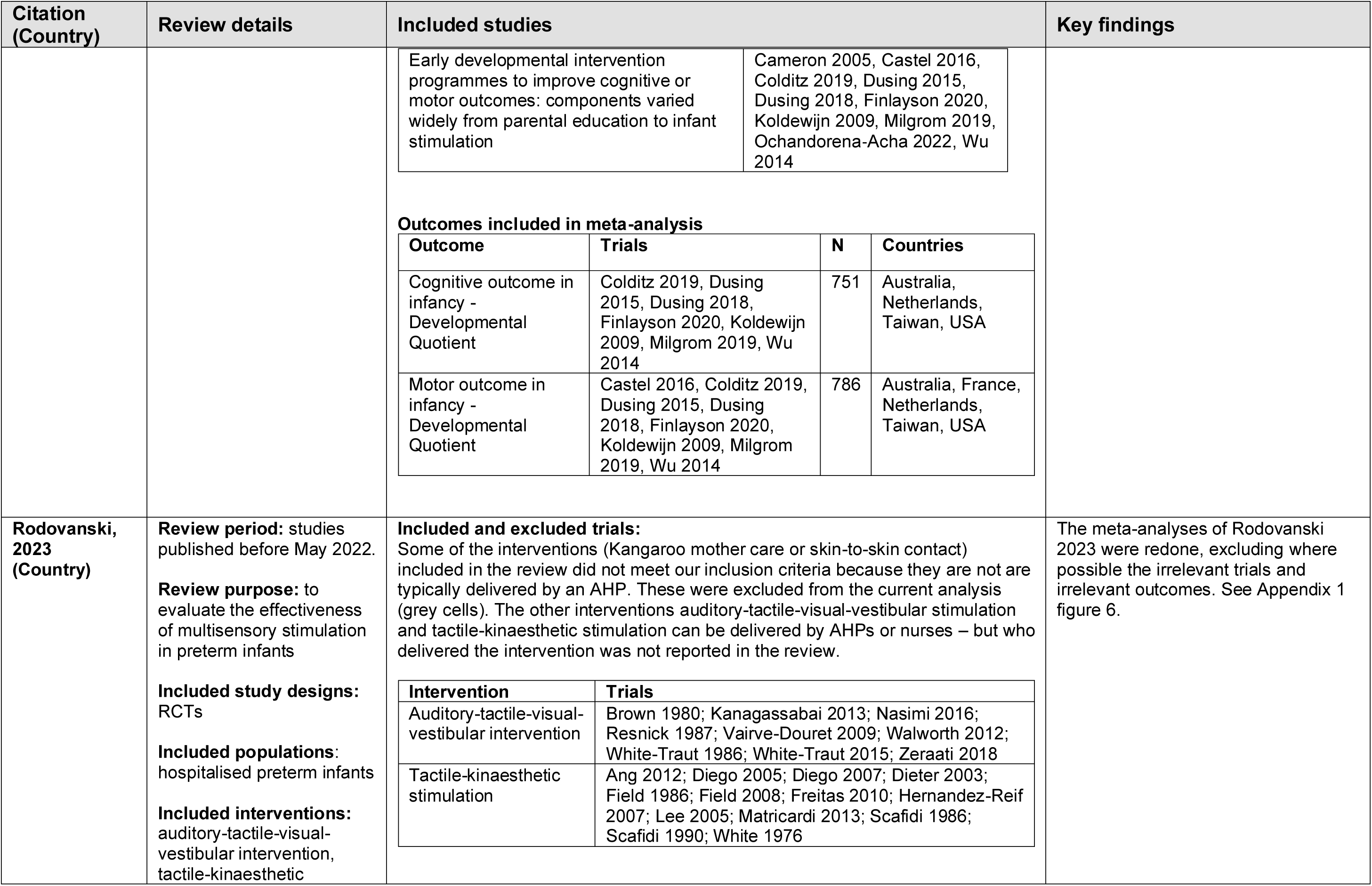
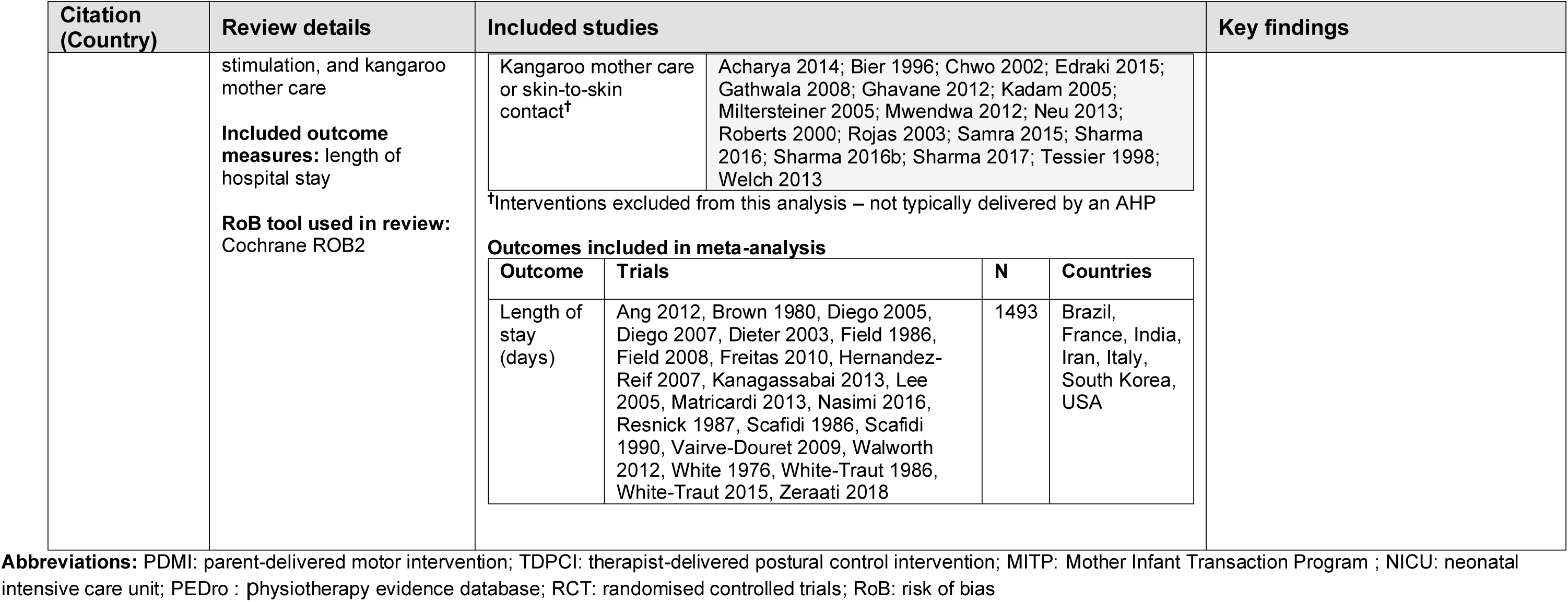
Summary of included studies for Q2.

All studies included for question 2 were systematic reviews.

### 6.3 Quality appraisal

#### Summary quality appraisal tables

**Table 10:** Quality appraisal results for cohort studies.

| Study | ROBINS-I domain level risk of bias judgements |  |  |  |  |  |  | Overall risk of bias |
| --- | --- | --- | --- | --- | --- | --- | --- | --- |
|  | Bias due to confounding | Bias in selection of participants into the study | Bias in classification of interventions | Bias due to deviations from intended interventions | Bias due to missing data | Bias in measurement of outcomes | Bias in selection of the reported result |  |
| Gover 2014 | Unclear (confounders adjusted for in analysis, but reporting unclear) | Low | Moderate (flexible definition of MDT) | Low | Low | Low | No information (no protocol) | Serious |

**Table 11:**
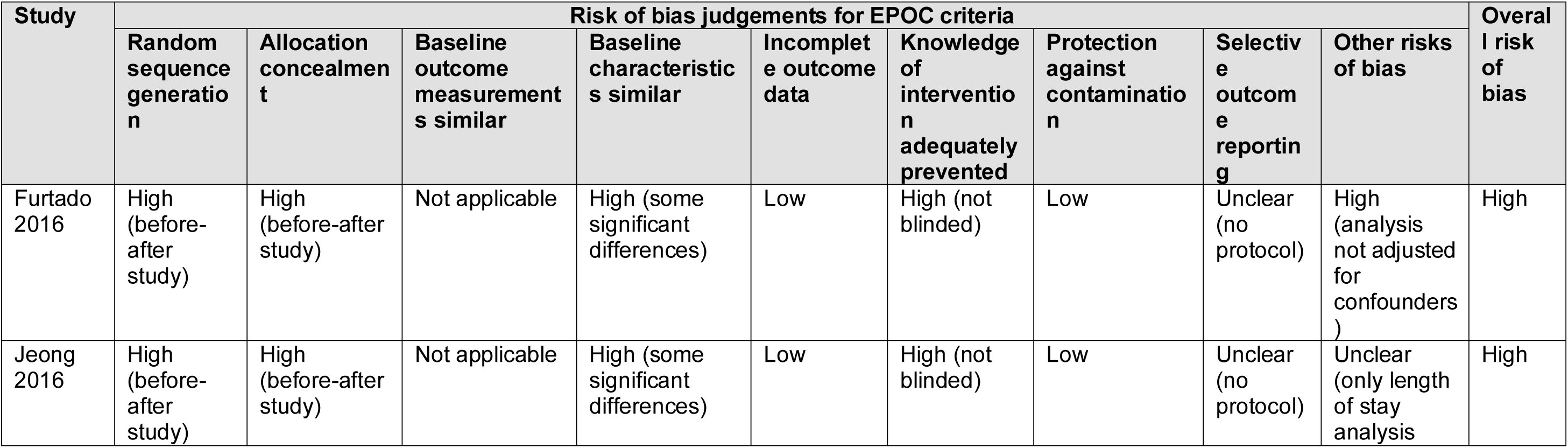
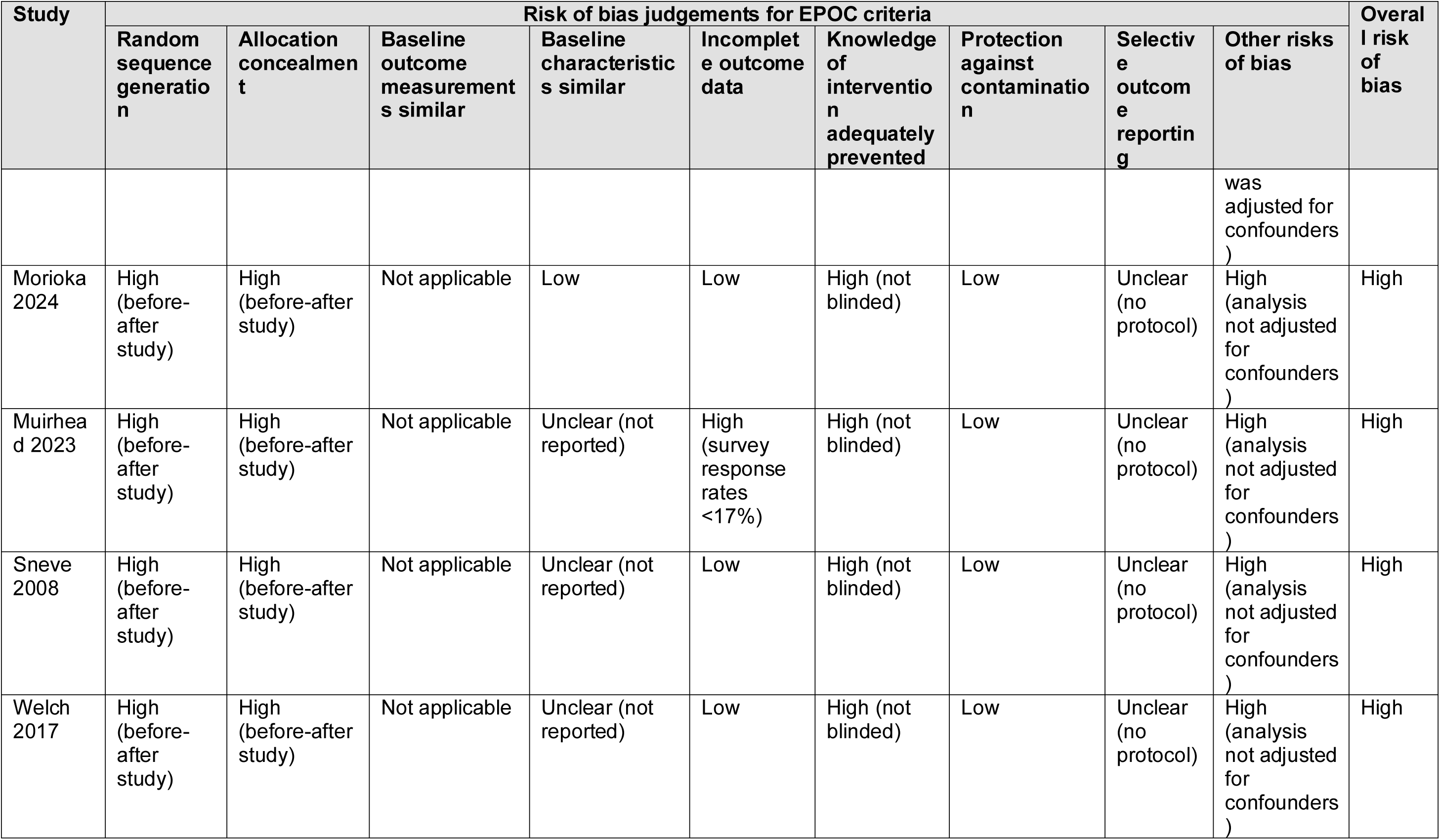
Quality appraisal results for before/after studies.

**Table 12:** Quality appraisal results for systematic reviews.

| Study | ROBIS domain judgements |  |  |  |  |
| --- | --- | --- | --- | --- | --- |
|  | Study eligibility criteria | Identification and selection of studies | Data collection and study appraisal | Synthesis and findings | Risk of bias in the review |
| Girabent-Farres 2021 | Low | Unclear (search terms simplistic) | Low | Unclear (cannot access protocol to check planned analyses) | Unclear |
| Greene 2023 | Low | Low | Low | Low | Low |
| Lavallee 2021 | Low | Low | Low | Low | Low |
| Orton 2024 | Low | Low | Low | Low | Low |
| Rodovanski 2023 | Low | Low | Low | Unclear (no pre-registered protocol to check planned analyses) | Unclear |

### 6.4 GRADE profiles

**Table 13:**
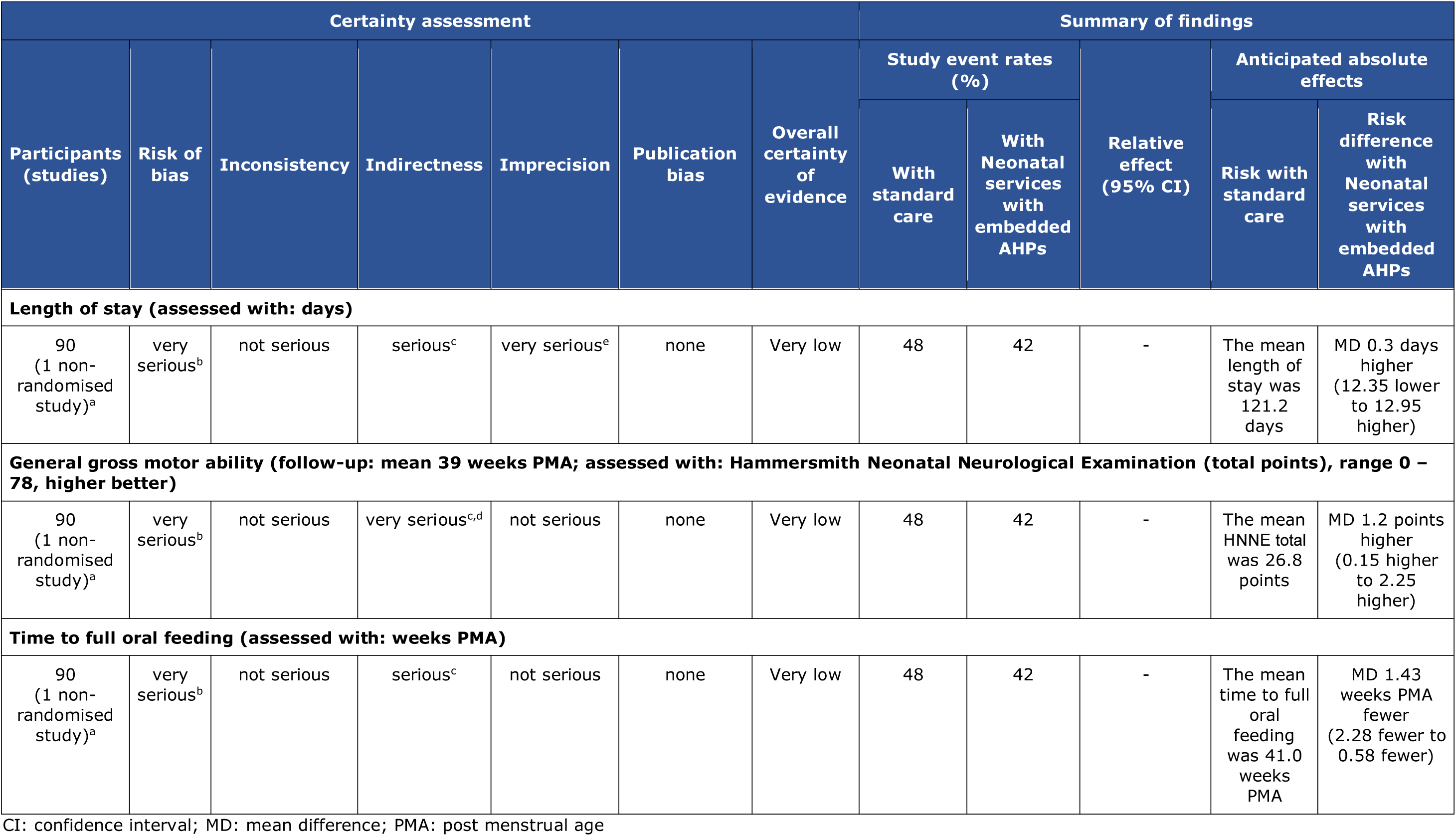
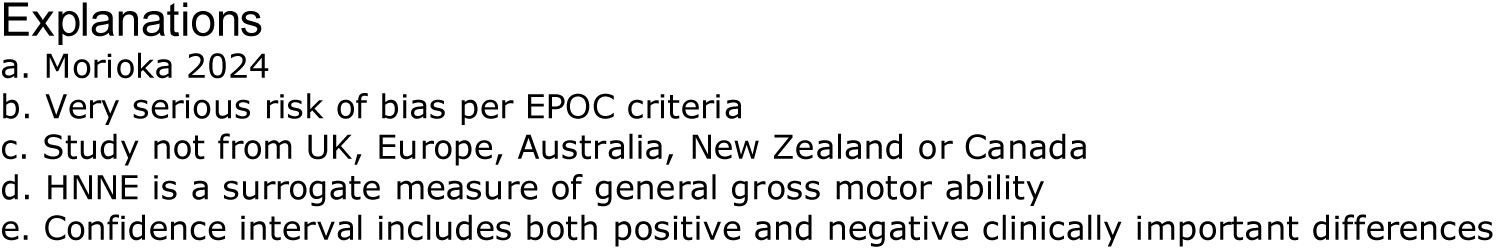
GRADE profile for Q1 - neonatal services with embedded allied health professionals.

| Certainty assessment |  |  |  |  |  |  | Summary of findings |  |  |  |  |
| --- | --- | --- | --- | --- | --- | --- | --- | --- | --- | --- | --- |
| Participants (studies) | Risk of bias | Inconsistency | Indirectness | Imprecision | Publication bias | Overall certainty of evidence | Study event rates (%) |  | Relative effect (95% CI) | Anticipated absolute effects |  |
|  |  |  |  |  |  |  | With standard care | With Neonatal services with embedded AHPs |  | Risk with standard care | Risk difference with Neonatal services with embedded AHPs |
| Length of stay (assessed with: days) |  |  |  |  |  |  |  |  |  |  |  |
| 90 (1 non-randomised study)<sup>a</sup> | very serious<sup>b</sup> | not serious | serious<sup>c</sup> | very serious<sup>e</sup> | none | Very low | 48 | 42 | - | The mean length of stay was 121.2 days | MD 0.3 days higher (12.35 lower to 12.95 higher) |
| General gross motor ability (follow-up: mean 39 weeks PMA; assessed with: Hammersmith Neonatal Neurological Examination (total points), range 0 – 78, higher better) |  |  |  |  |  |  |  |  |  |  |  |
| 90 (1 non-randomised study)<sup>a</sup> | very serious<sup>b</sup> | not serious | very serious<sup>c,d</sup> | not serious | none | Very low | 48 | 42 | - | The mean HNNE total was 26.8 points | MD 1.2 points higher (0.15 higher to 2.25 higher) |
| Time to full oral feeding (assessed with: weeks PMA) |  |  |  |  |  |  |  |  |  |  |  |
| 90 (1 non-randomised study)<sup>a</sup> | very serious<sup>b</sup> | not serious | serious<sup>c</sup> | not serious | none | Very low | 48 | 42 | - | The mean time to full oral feeding was 41.0 weeks PMA | MD 1.43 weeks PMA fewer (2.28 fewer to 0.58 fewer) |

CI: confidence interval; MD: mean difference; PMA: post menstrual age

**Table 14:**
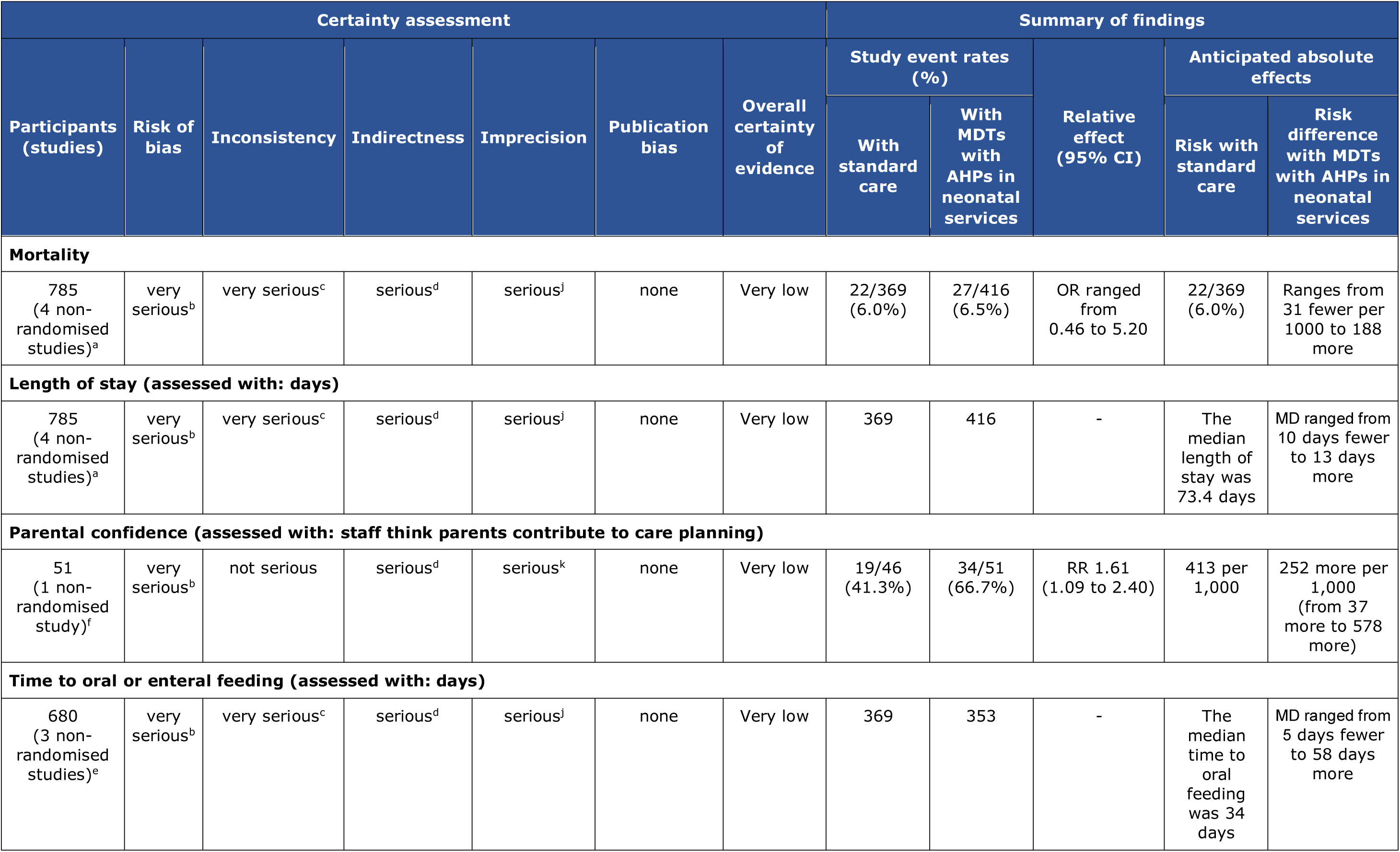
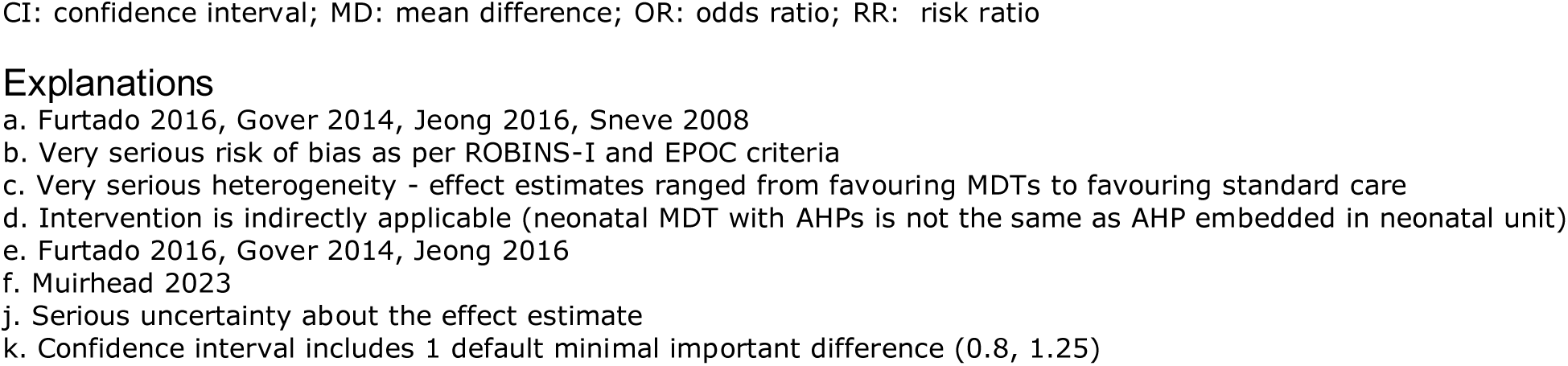
GRADE profile for Q1 – multidisciplinary teams with allied health professionals.

| Certainty assessment |  |  |  |  |  |  | Summary of findings |  |  |  |  |
| --- | --- | --- | --- | --- | --- | --- | --- | --- | --- | --- | --- |
| Participants (studies) | Risk of bias | Inconsistency | Indirectness | Imprecision | Publication bias | Overall certainty of evidence | Study event rates (%) |  | Relative effect (95% CI) | Anticipated absolute effects |  |
|  |  |  |  |  |  |  | With standard care | With MDTs with AHPs in neonatal services |  | Risk with standard care | Risk difference with MDTs with AHPs in neonatal services |
| Mortality |  |  |  |  |  |  |  |  |  |  |  |
| 785 (4 non-randomised studies)<sup>a</sup> | very serious<sup>b</sup> | very serious<sup>c</sup> | serious<sup>d</sup> | serious<sup>j</sup> | none | Very low | 22/369 (6.0%) | 27/416 (6.5%) | OR ranged from 0.46 to 5.20 | 22/369 (6.0%) | Ranges from 31 fewer per 1000 to 188 more |
| Length of stay (assessed with: days) |  |  |  |  |  |  |  |  |  |  |  |
| 785 (4 non-randomised studies)<sup>a</sup> | very serious<sup>b</sup> | very serious<sup>c</sup> | serious<sup>d</sup> | serious<sup>j</sup> | none | Very low | 369 | 416 | - | The median length of stay was 73.4 days | MD ranged from 10 days fewer to 13 days more |
| Parental confidence (assessed with: staff think parents contribute to care planning) |  |  |  |  |  |  |  |  |  |  |  |
| 51 (1 non-randomised study)<sup>f</sup> | very serious<sup>b</sup> | not serious | serious<sup>d</sup> | serious<sup>k</sup> | none | Very low | 19/46 (41.3%) | 34/51 (66.7%) | RR 1.61 (1.09 to 2.40) | 413 per 1,000 | 252 more per 1,000 (from 37 more to 578 more) |
| Time to oral or enteral feeding (assessed with: days) |  |  |  |  |  |  |  |  |  |  |  |
| 680 (3 non-randomised studies)<sup>e</sup> | very serious<sup>b</sup> | very serious<sup>c</sup> | serious<sup>d</sup> | serious<sup>j</sup> | none | Very low | 369 | 353 | - | The median time to oral feeding was 34 days | MD ranged from 5 days fewer to 58 days more |

CI: confidence interval; MD: mean difference; OR: odds ratio; RR: risk ratio

**Table 15:**
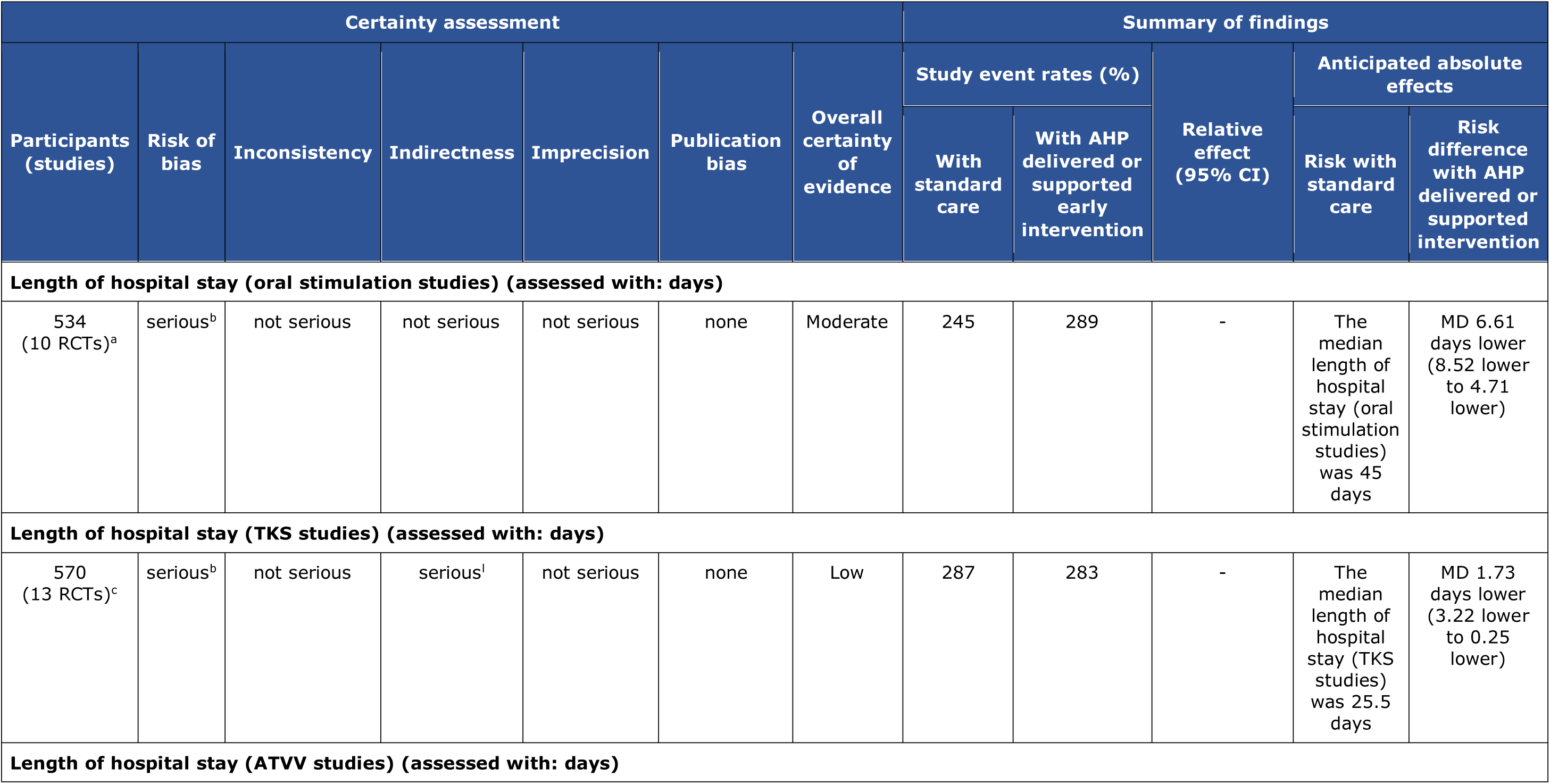
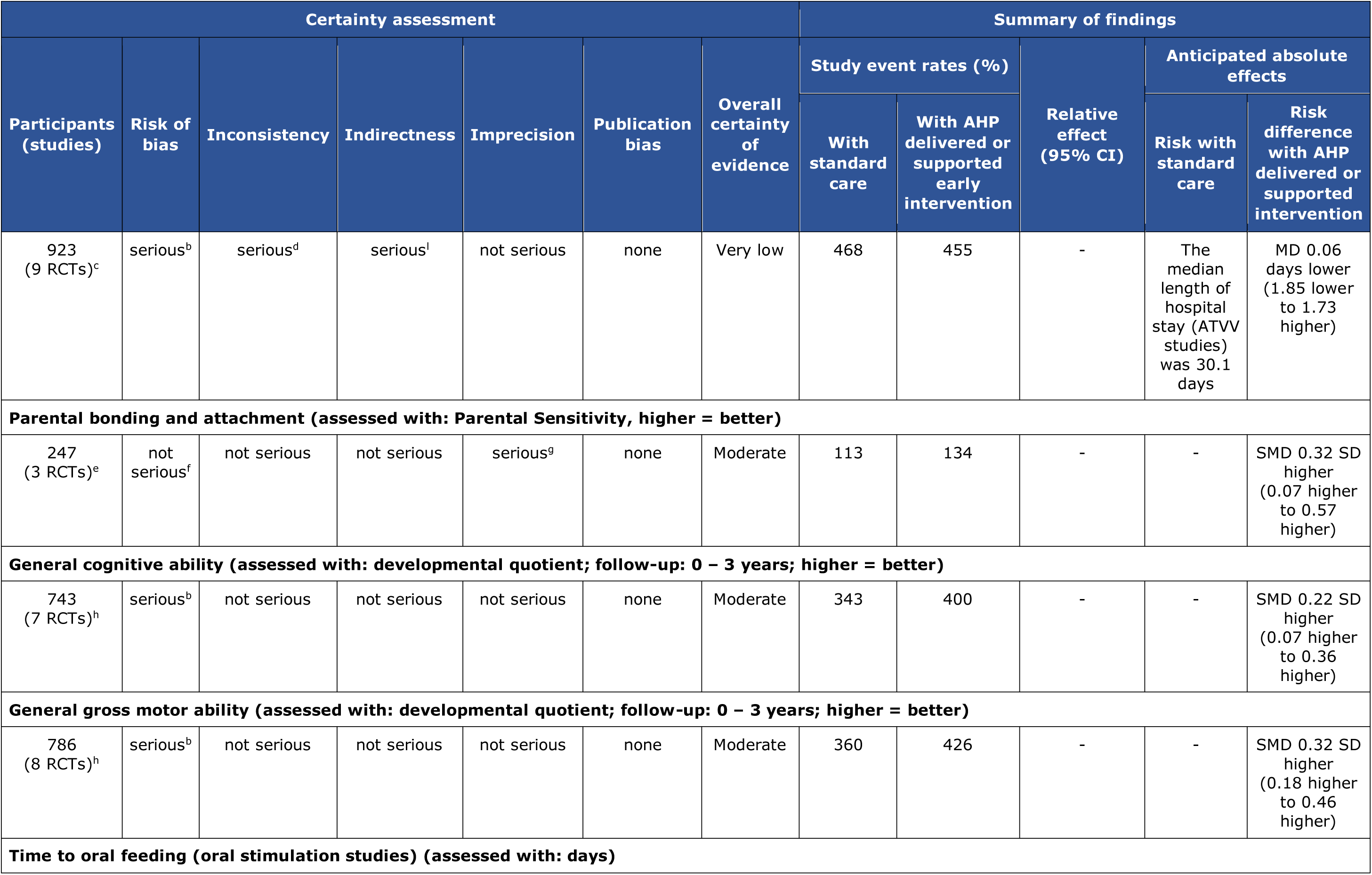
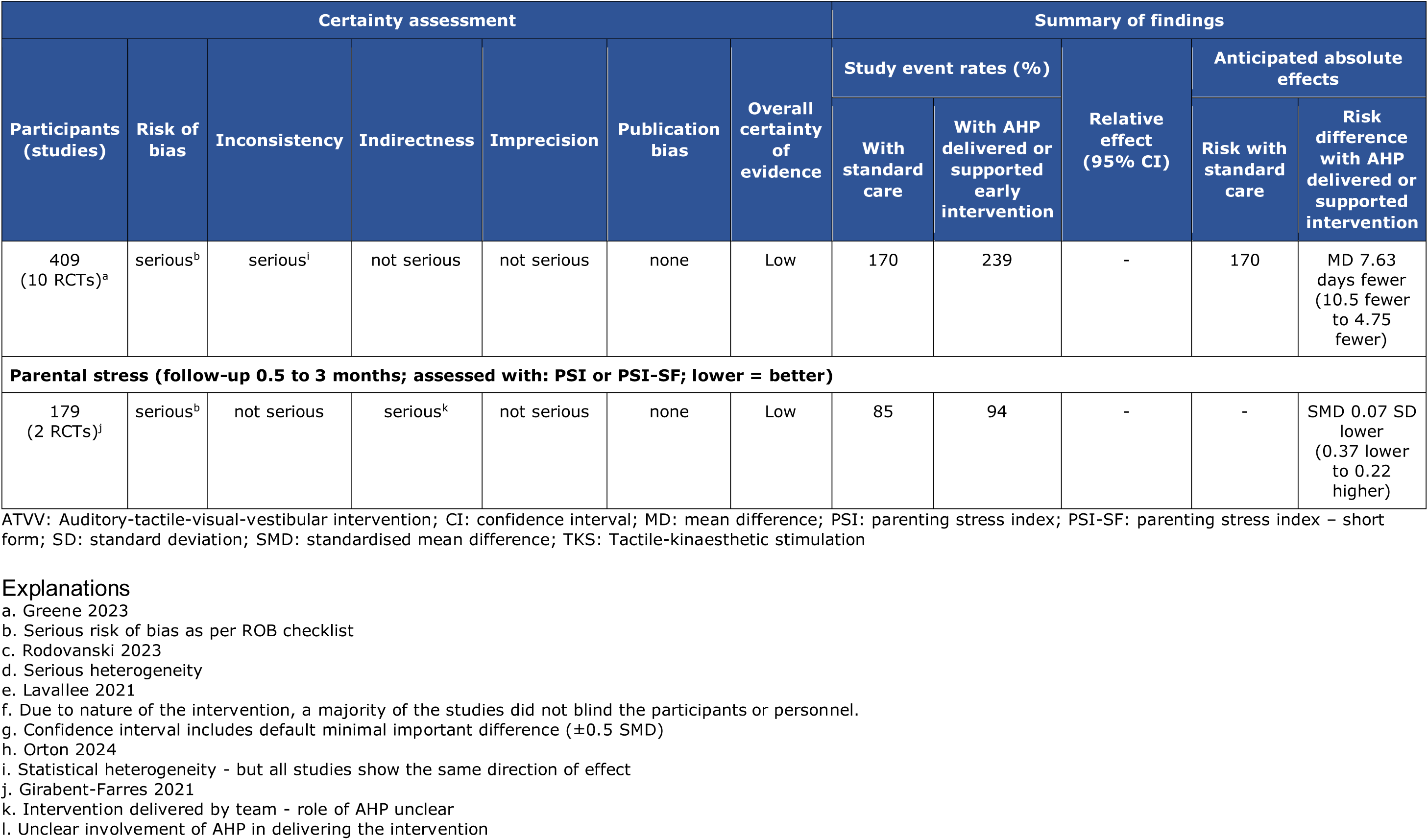
GRADE profile for Q2 - early interventions provided by allied health professionals in neonatal units.

### 6.5 Information available on request

Protocol, search strategies, and excluded studies.

## Acknowledgements

The authors would like to thank Amanda Lawes, Beti-jane Ingram, Bethan Phillips, Catherine Pape, Catriona Matthews, Ceri Selman, Debbie Paris, Elen Elias, Fiona Luff, Leah Watson, Louise Leach, Margaret Manton, and Stephanie Griffiths for their expertise in guiding the review question, and helpful comments on draft versions of the review protocol and rapid review report.

## 8. APPENDICES

## 8.1 APPENDIX 1: Forest plots

**Forest plots for review question 1: What is the effectiveness of neonatal services with embedded AHPs compared to neonatal services without embedded AHPs?**

**Figure 2.**
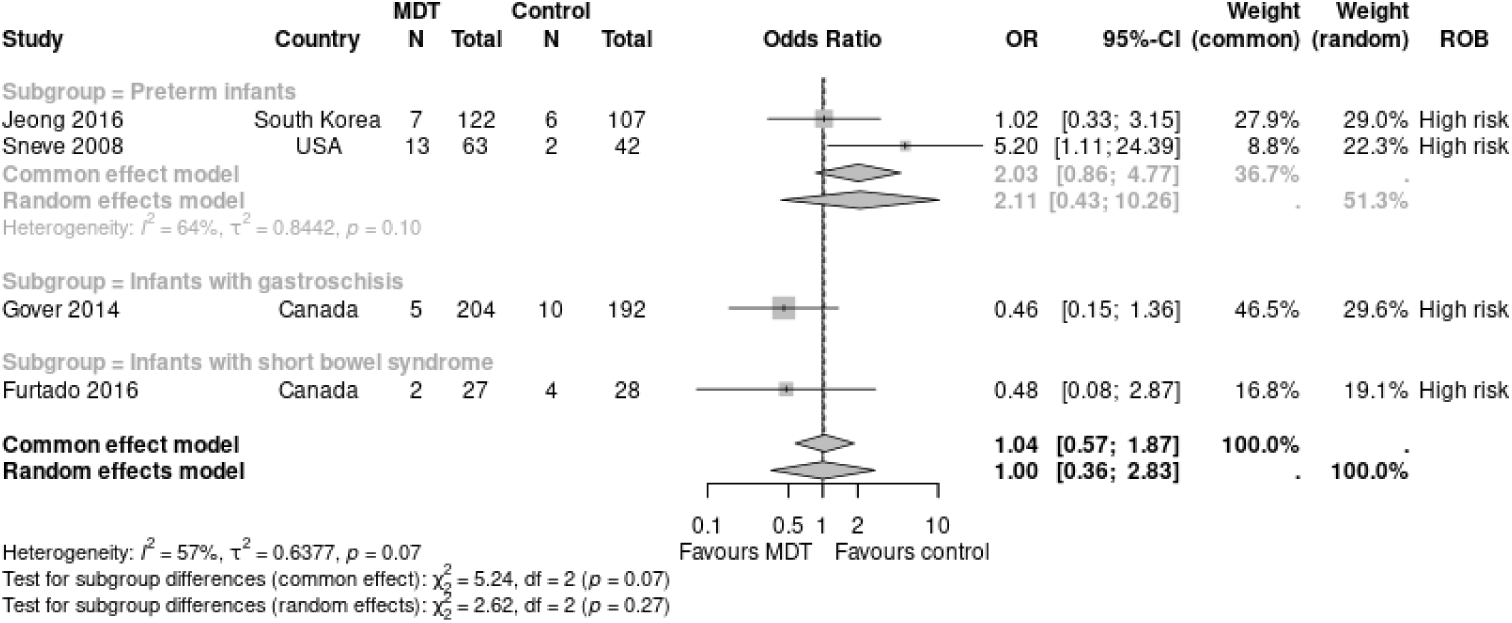
After versus before implementation of an MDT with a dietician: mortality. AHP: allied health professional; CI: confidence interval; OR: odds ratio; MDT: multidisciplinary team; ROB: risk of bias

**Figure 3.**
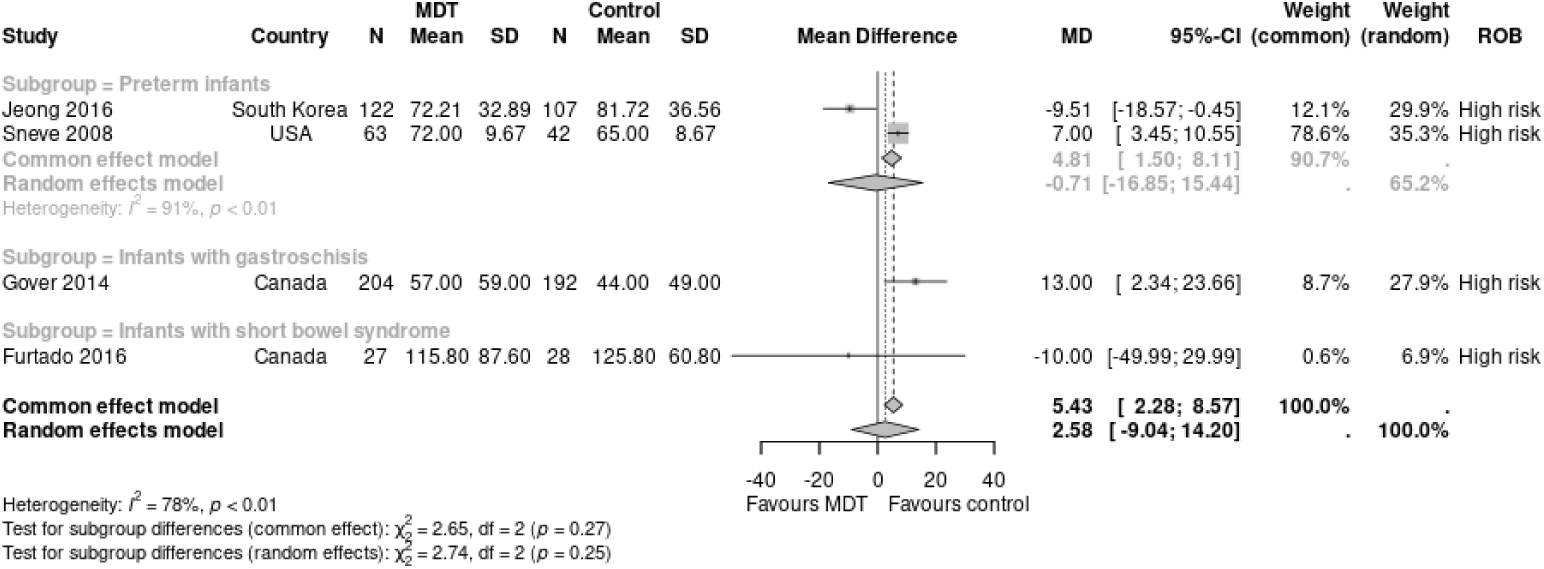
After versus before implementation of an MDT with a dietician: length of NICU stay (days) AHP: allied health professional; CI: confidence interval; MD: mean difference; MDT: multidisciplinary team; ROB: risk of bias; SD: standard deviation

**Figure 4.**
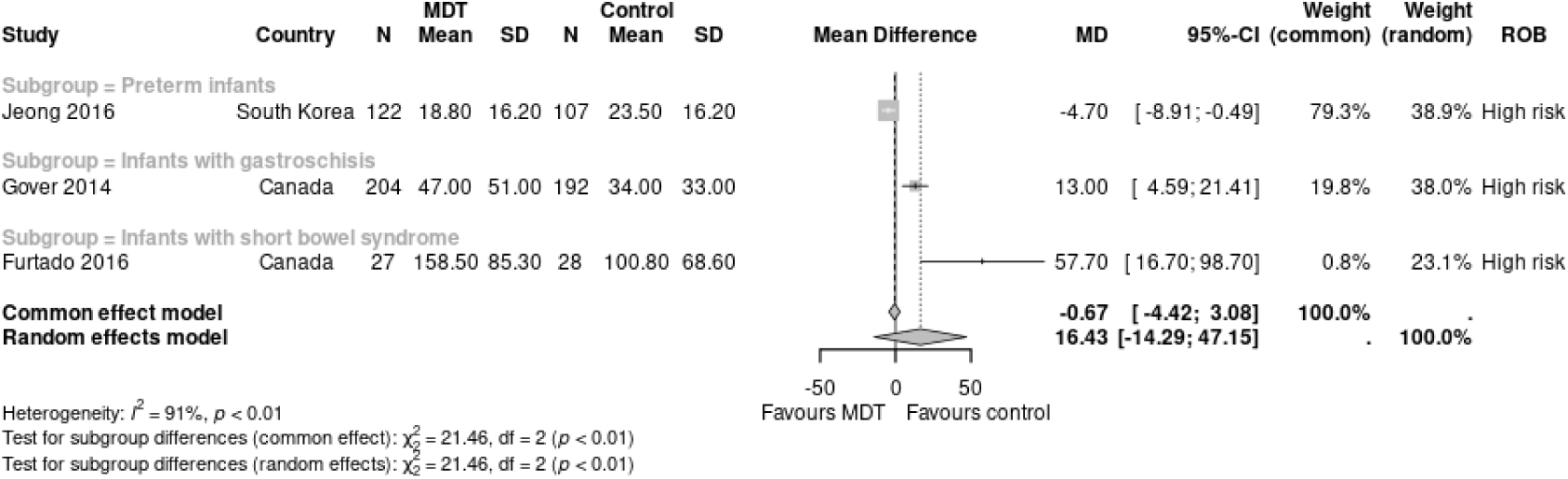
After versus before implementation of an MDT with a dietician: time to oral or enteral feeding (days) AHP: allied health professional; CI: confidence interval; MD: mean difference; MDT: multidisciplinary team; ROB: risk of bias; SD: standard deviation

**Forest plots for review question 2: What is the effectiveness of early interventions provided by AHPs in neonatal units?**

**Figure 5.**
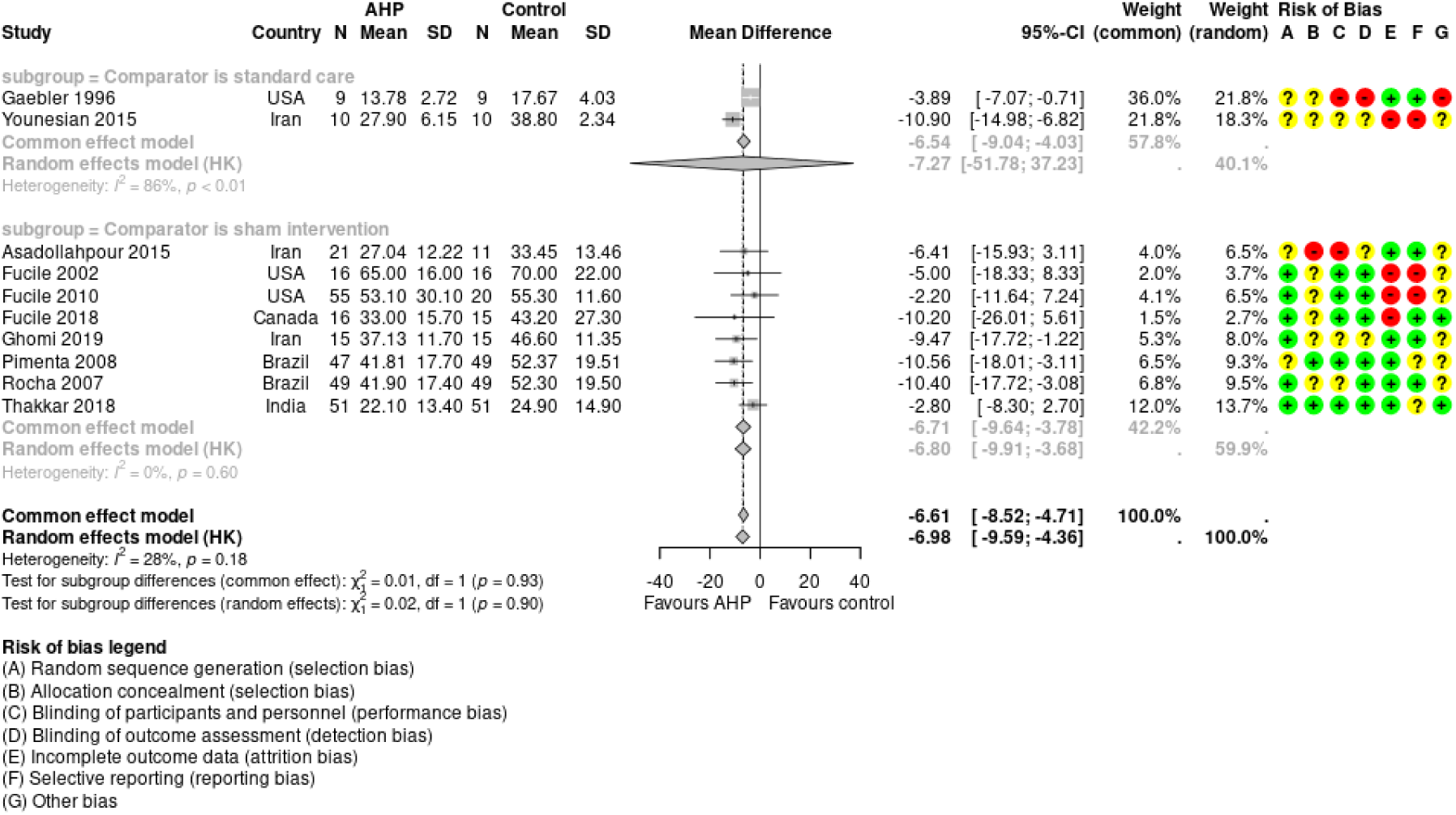
Early intervention (oral stimulation) provided by AHP versus standard care or sham stimulation: length of stay (days) AHP: allied health professional; CI: confidence interval; MD: mean difference; SD: standard deviation

**Figure 6.**
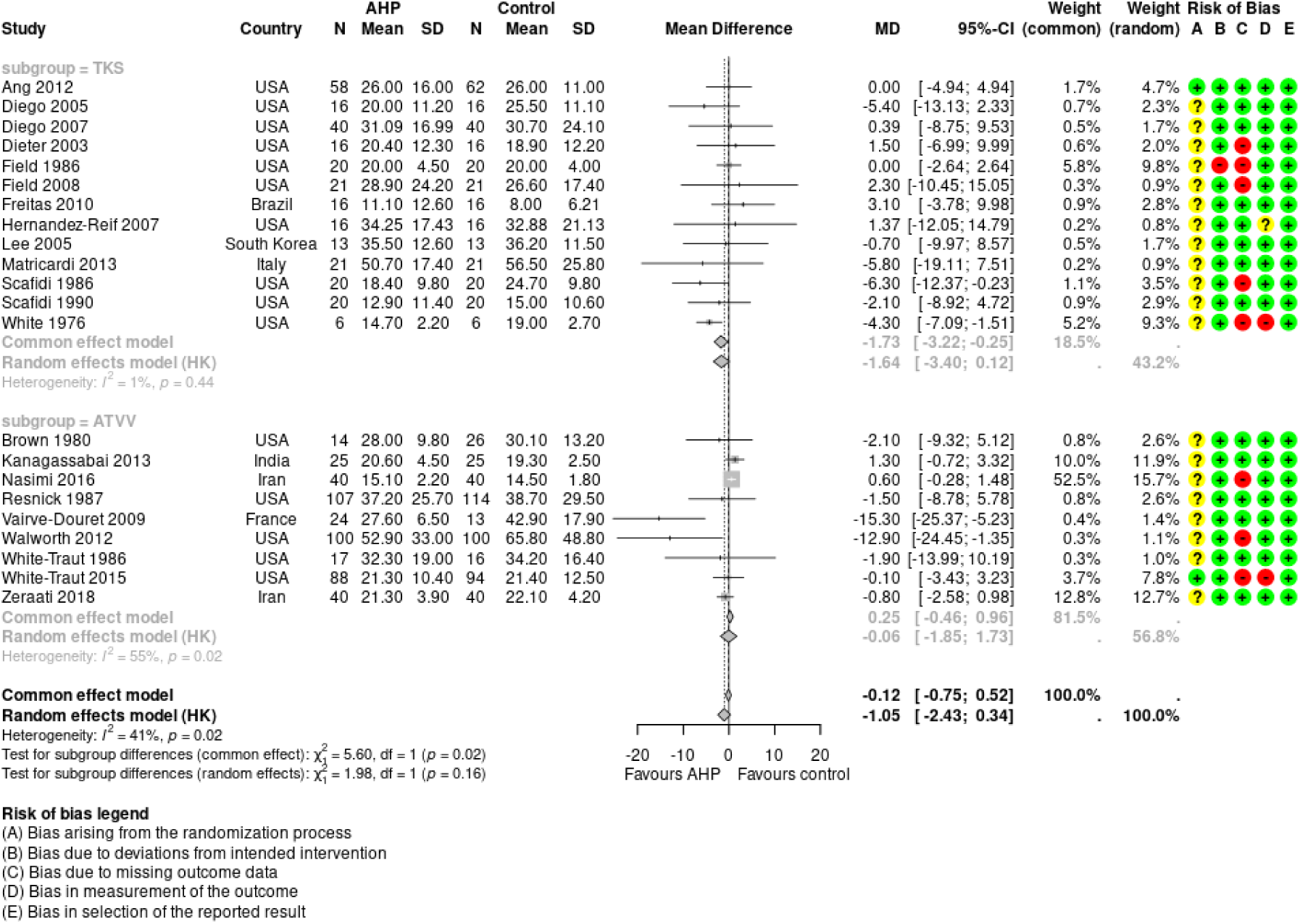
Early intervention (multisensory stimulation) provided by AHP versus standard care: length of stay (days) AHP: allied health professional; ATVV: auditory-tactile-visual-vestibular intervention; CI: confidence interval; MD: mean difference; SD: standard deviation; TKS: tactile-kinaesthetic stimulation

**Figure 7.**
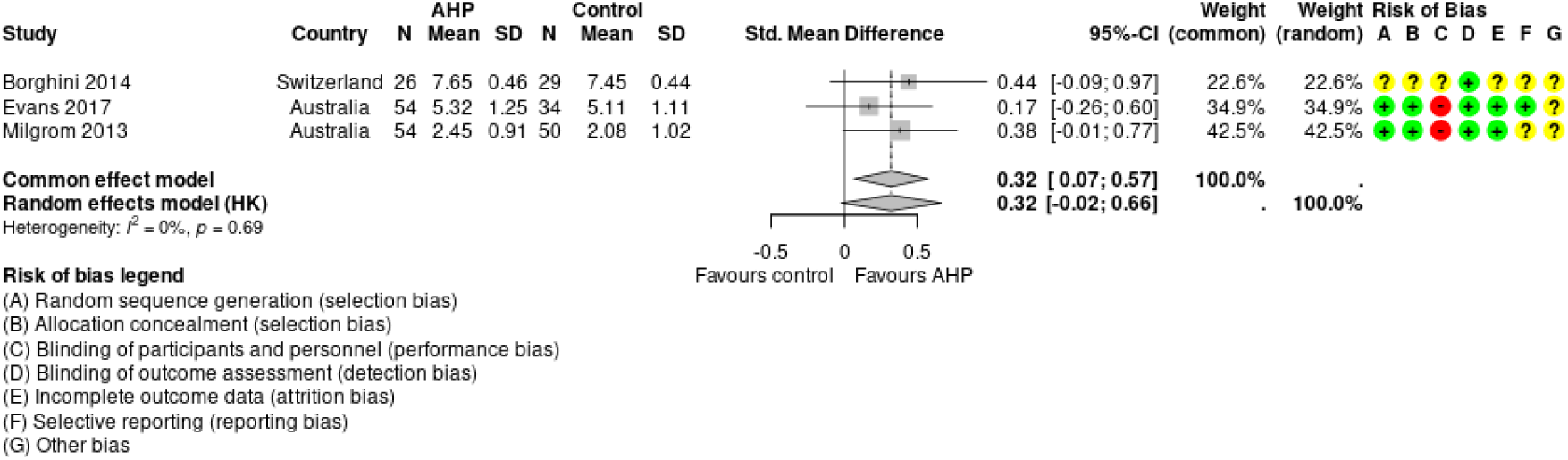
Early interventions provided by AHP for parents versus standard care. Parental bonding and attachment: measured with parental sensitivity (higher = better) AHP: allied health professional; CI: confidence interval; SD: standard deviation; SMD: standardized mean difference

**Figure 8.**
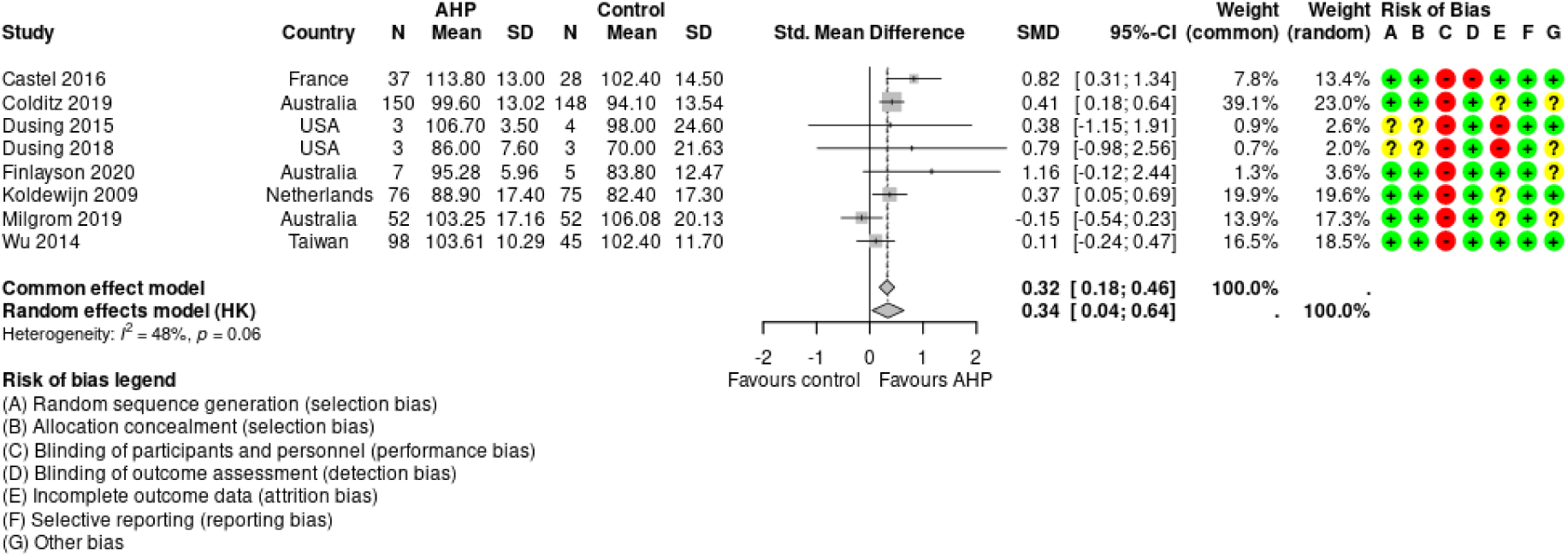
Early interventions provided by AHP versus standard care. General gross motor ability measured with developmental quotient in infancy (higher = better) AHP: allied health professional; CI: confidence interval; SD: standard deviation; SMD: standardized mean difference

**Figure 9.**
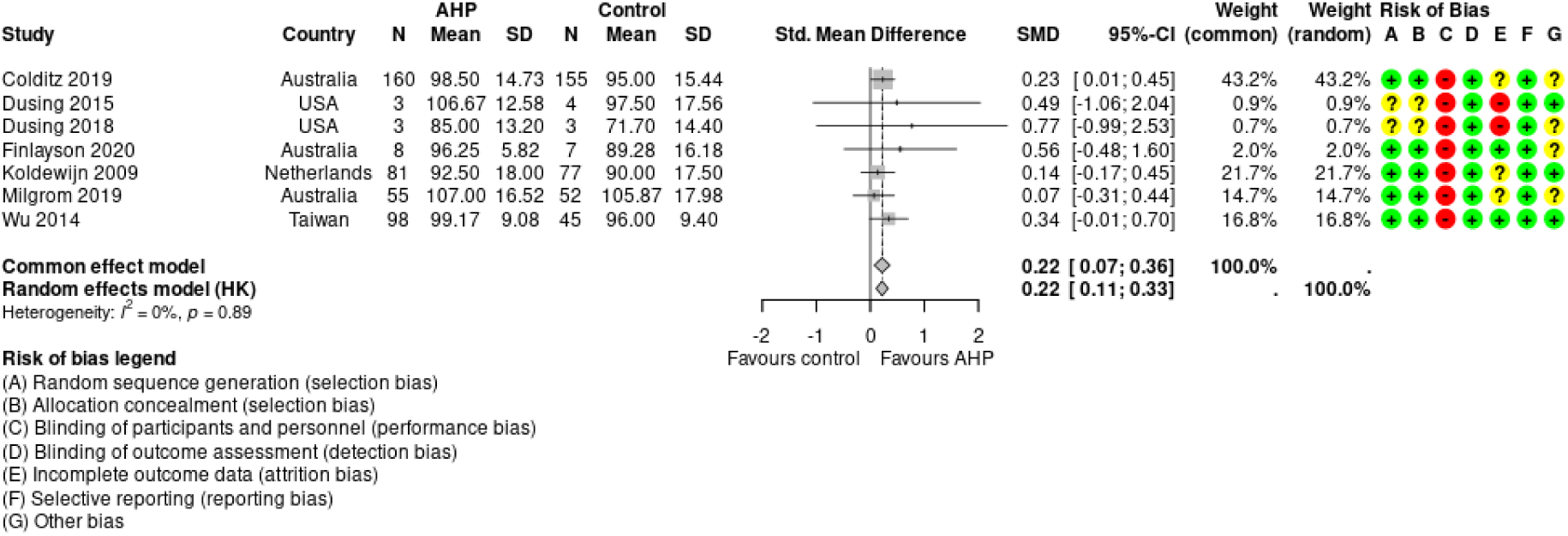
Early interventions provided by AHP versus standard care. General cognitive ability measured with developmental quotient in infancy (higher = better) AHP: allied health professional; CI: confidence interval; SD: standard deviation; SMD: standardized mean difference

**Figure 10.**
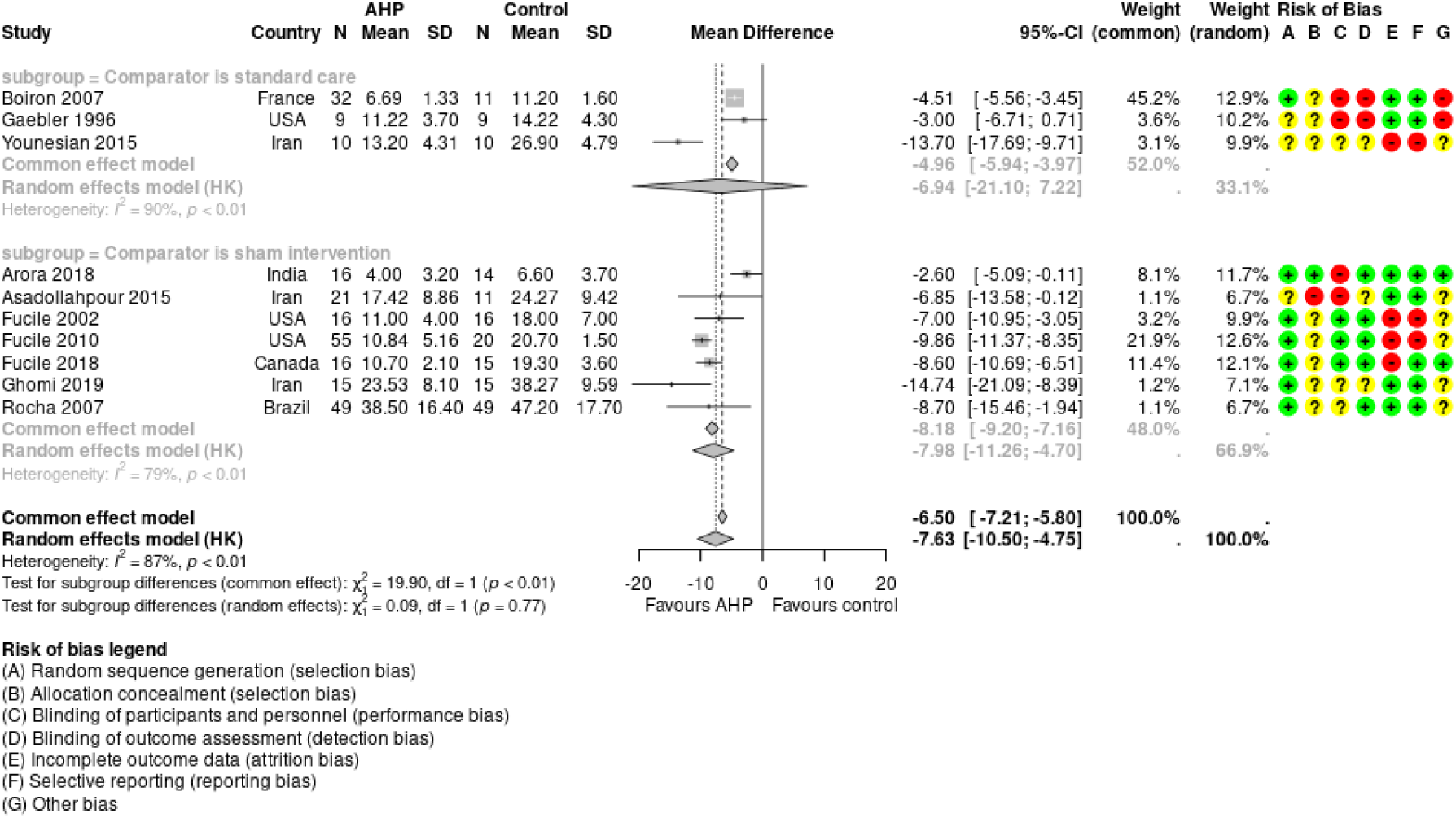
Oral stimulation provided by AHP versus standard care or sham intervention. Oral feeding measured by time to oral feeding (days) AHP: allied health professional; CI: confidence interval; MD: mean difference; SD: standard deviation

**Figure 11.**
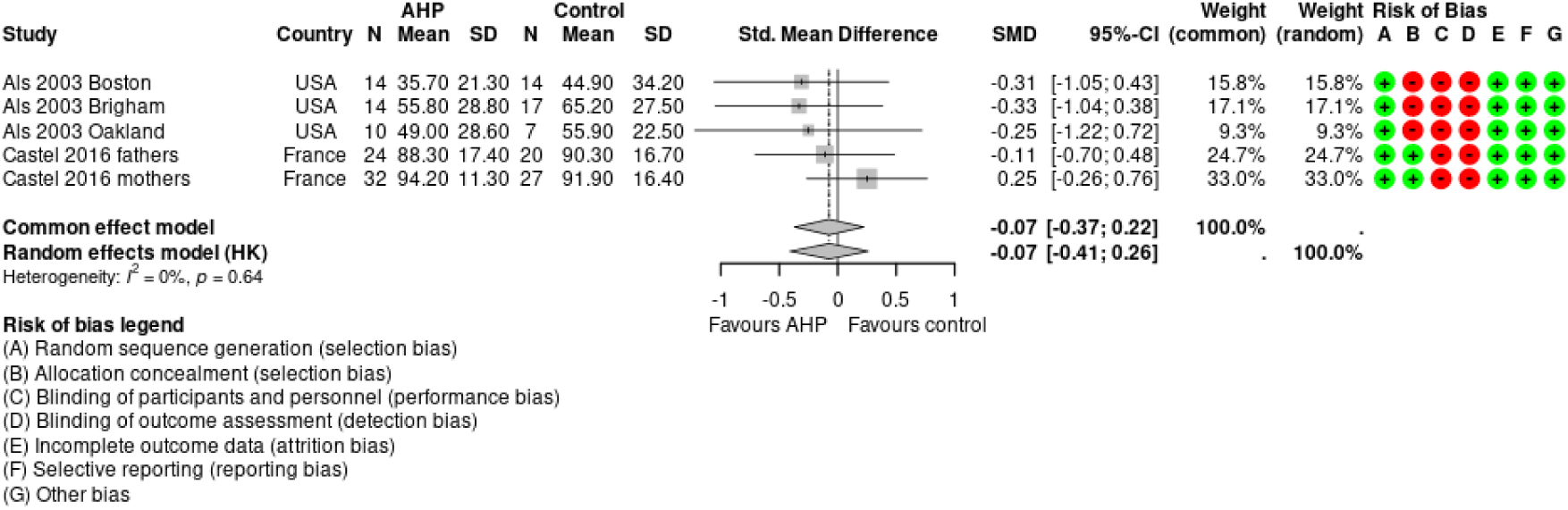
Early intervention provided by AHP versus standard care. Parental mental health and mood measured by parental stress (lower = better) AHP: allied health professional; CI: confidence interval; SD: standard deviation; SMD: standardized mean difference

## 8.2 APPENDIX 2: Search strategy

**Table 16.** Summary of searches.

| Database | Search 1 | Search 2 | Overlap |
| --- | --- | --- | --- |
| Emcare | 525 | 1206 | 14 |
| Medline | 428 | (4208 (2039 SRS, 2669 RCTS)) | NA |
| AMED | 39 | 25 | 3 |
| CINAHL | 316 | 1628 | 14 |
| Central | 188 | 1446 | 31 |

**Table 17.**
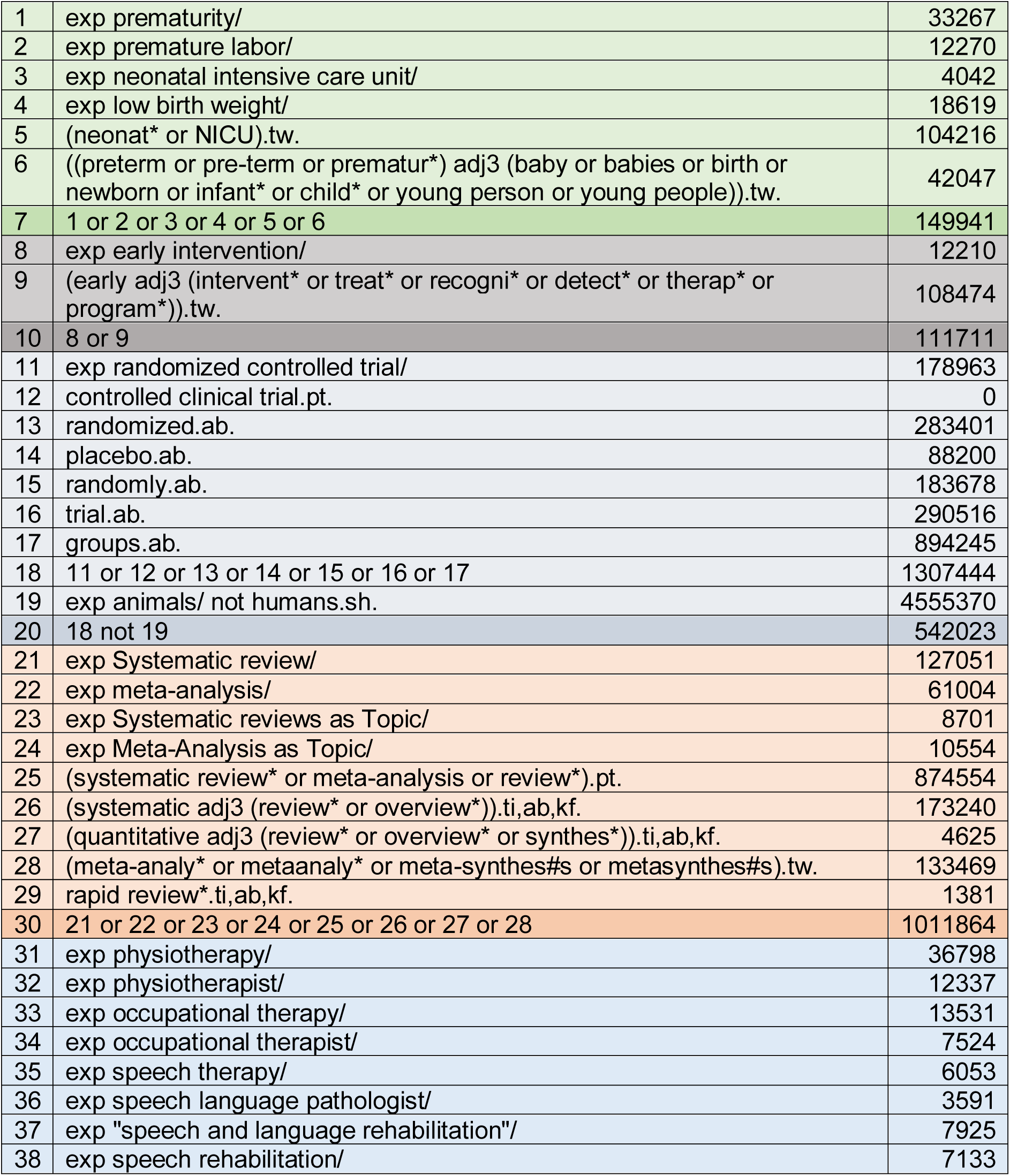
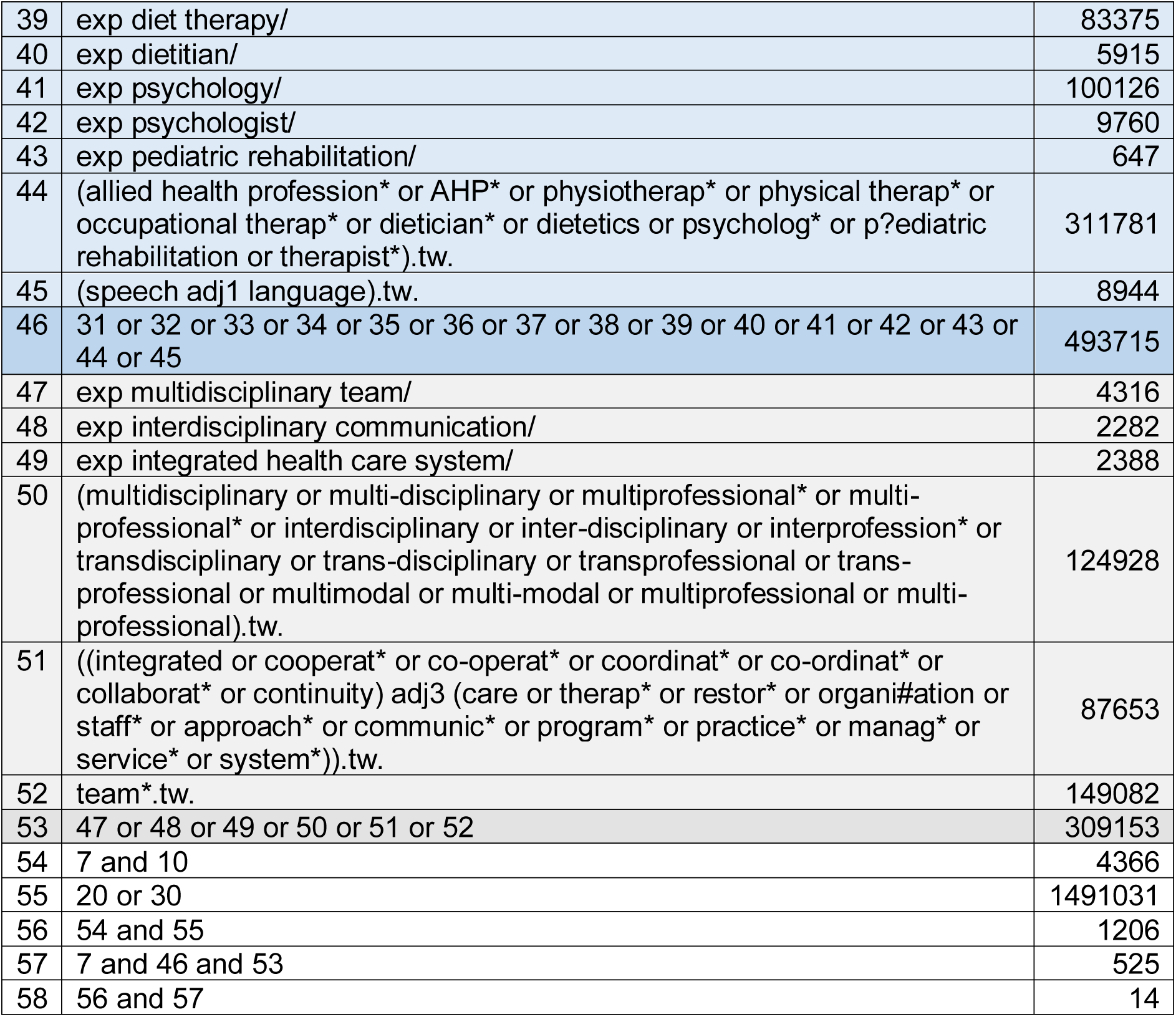
Ovid Emcare <1995 to 2024 Week 09>.

**Table 18.**
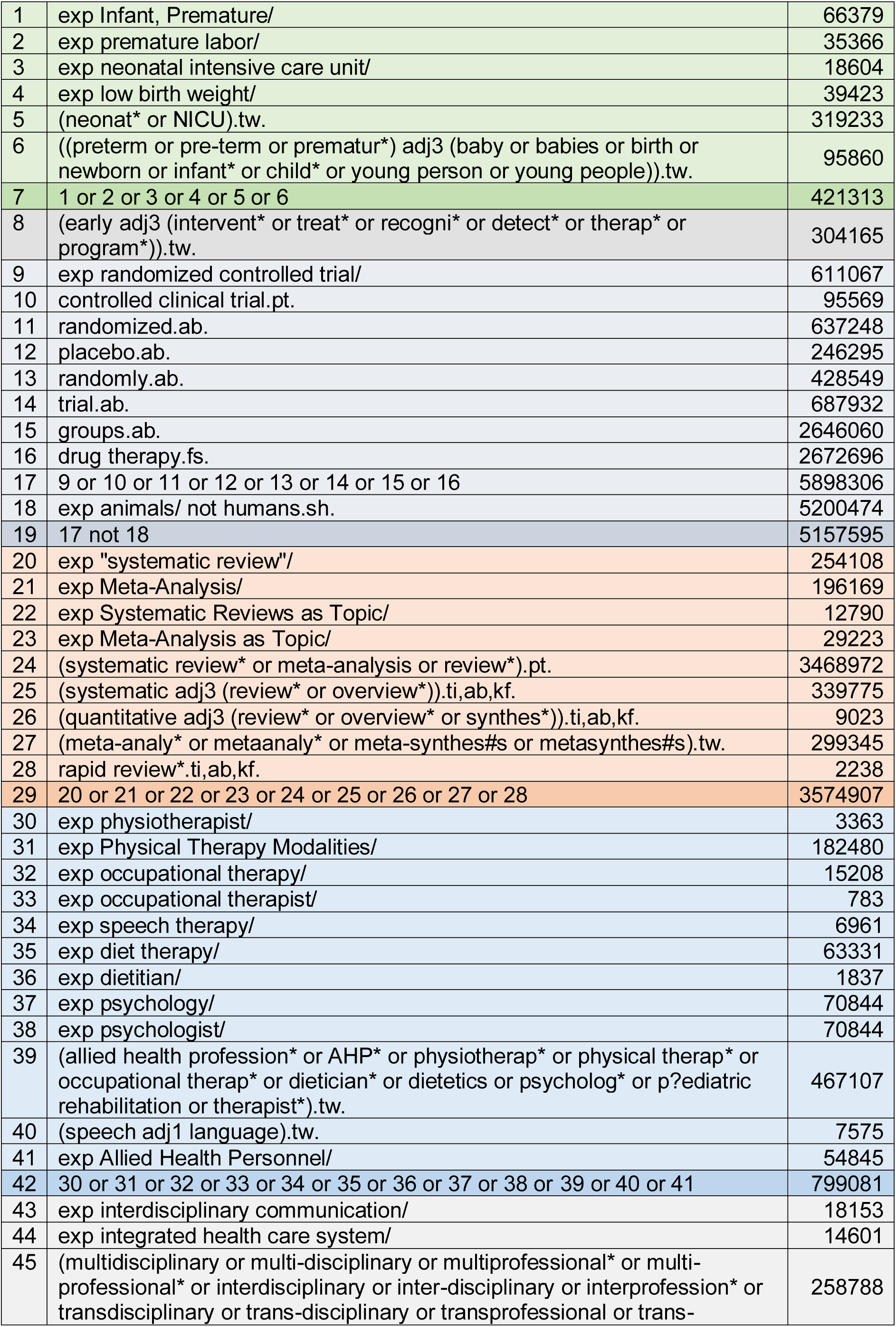
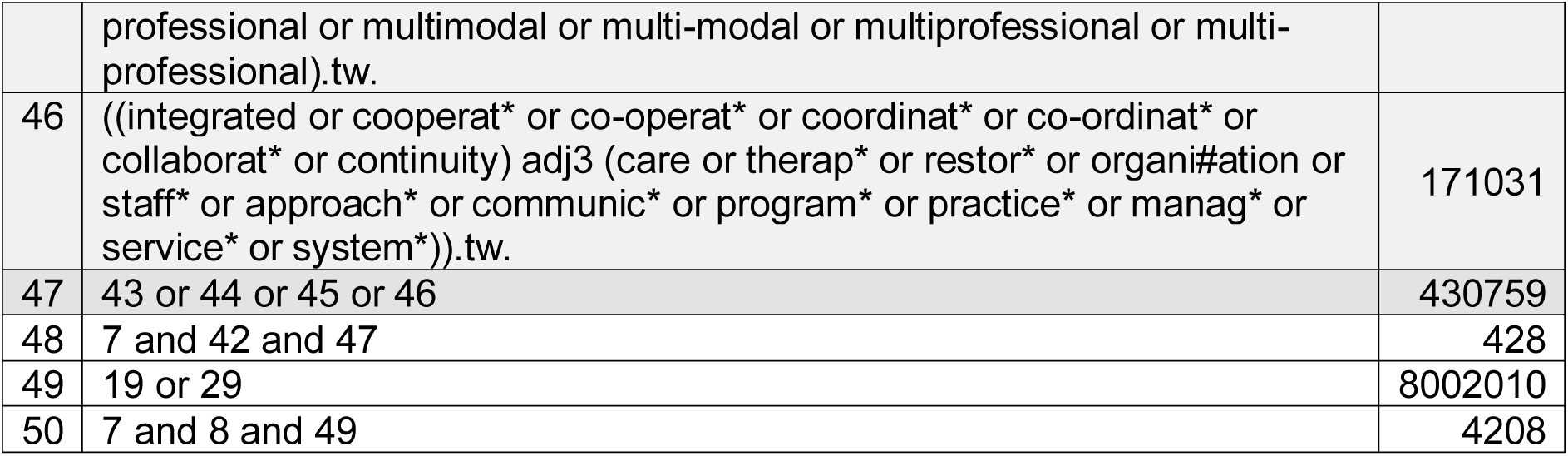
Ovid MEDLINE(R) ALL <1946 to March 06, 2024>.

**Table 19.** AMED (Allied and Complementary Medicine) <1985 to October 2023>.

|  |  |  |
| --- | --- | --- |
| 1 | exp Infant, Premature/ | 212 |
| 2 | exp intensive care neonatal/ | 78 |
| 3 | exp infant newborn/ or infant low birth weight/ or infant premature/ | 802 |
| 4 | (neonat* or NICU).tw. | 646 |
| 5 | ((preterm or pre-term or prematur*) adj3 (baby or babies or birth or newborn or infant* or child* or young person or young people)).tw. | 420 |
| 6 | 1 or 2 or 3 or 4 or 5 | 1308 |
| 7 | (early adj3 (intervent* or treat* or recogni* or detect* or therap* or program*)).tw. | 2638 |
| 8 | exp randomized controlled trial/ | 3211 |
| 9 | controlled clinical trial.pt. | 70 |
| 10 | randomized.ab. | 12648 |
| 11 | placebo.ab. | 3575 |
| 12 | randomly.ab. | 8323 |
| 13 | trial.ab. | 11221 |
| 14 | groups.ab. | 31019 |
| 15 | 8 or 9 or 10 or 11 or 12 or 13 or 14 | 46002 |
| 16 | exp animals/ not humans.sh. | 16334 |
| 17 | 15 not 16 | 42827 |
| 18 | exp meta-analysis/ | 427 |
| 19 | (systematic review* or meta-analysis or review*).pt. | 14041 |
| 20 | (systematic adj3 (review* or overview*)).ti,ab. | 7324 |
| 21 | (quantitative adj3 (review* or overview* or synthes*)).ti,ab. | 210 |
| 22 | (meta-analy* or metaanaly* or meta-synthes#s or metasynthes#s).tw. | 3925 |
| 23 | rapid review*.ti,ab. | 40 |
| 24 | 18 or 19 or 20 or 21 or 22 or 23 | 16824 |
| 25 | exp Physical Therapy Modalities/ | 32670 |
| 26 | exp Physiotherapists/ | 1713 |
| 27 | exp occupational therapists/ | 1197 |
| 28 | exp Speech therapy/ | 1143 |
| 29 | exp diet therapy/ | 2229 |
| 30 | exp psychology/ | 37093 |
| 31 | (allied health profession* or AHP* or physiotherap* or physical therap* or occupational therap* or dietician* or dietetics or psycholog* or p?ediatric rehabilitation or therapist*).tw. | 83670 |
| 32 | (speech adj1 language).tw. | 1003 |
| 33 | exp Allied Health Personnel/ | 693 |
| 34 | 25 or 26 or 27 or 28 or 29 or 30 or 31 or 32 or 33 | 106315 |
| 35 | exp patient care team/ | 1829 |
| 36 | (multidisciplinary or multi-disciplinary or multiprofessional* or multi-professional* or interdisciplinary or inter-disciplinary or interprofession* or transdisciplinary or trans-disciplinary or transprofessional or trans-professional or multimodal or multi-modal or multiprofessional or multi-professional).tw. | 6294 |
| 37 | ((integrated or cooperat* or co-operat* or coordinat* or co-ordinat* or collaborat* or continuity) adj3 (care or therap* or restor* or organi#ation or staff* or approach* or communic* or program* or practice* or manag* or service* or system*)).tw. | 4224 |
| 38 | team*.tw. | 6675 |
| 39 | 35 or 36 or 37 or 38 | 14025 |
| 40 | 6 and 34 and 39 | 39 |
| 41 | 17 or 24 | 55513 |
| 42 | 6 and 7 and 41 | 25 |
| 43 | 40 and 42 | 3 |

**Table 20.**
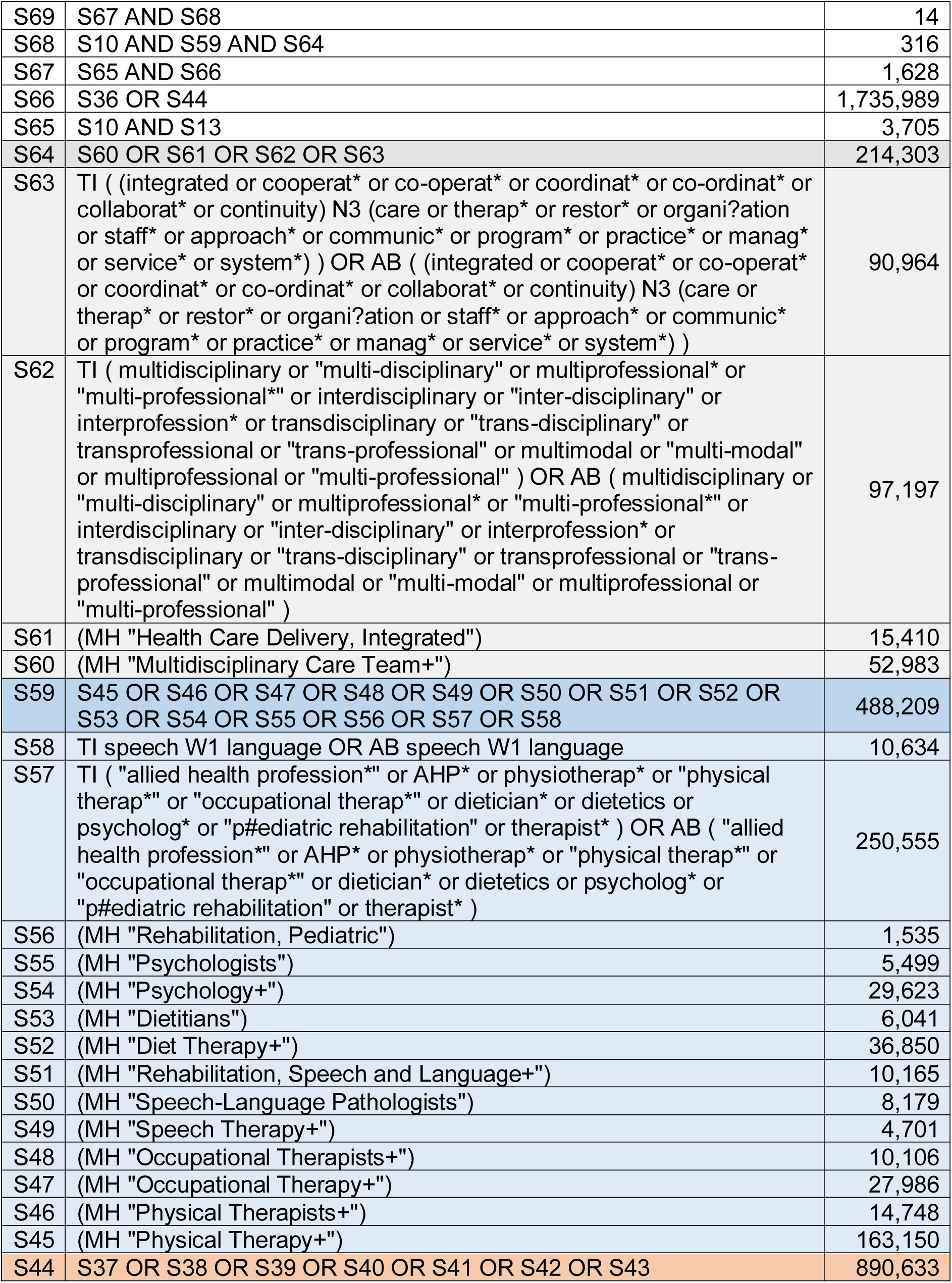
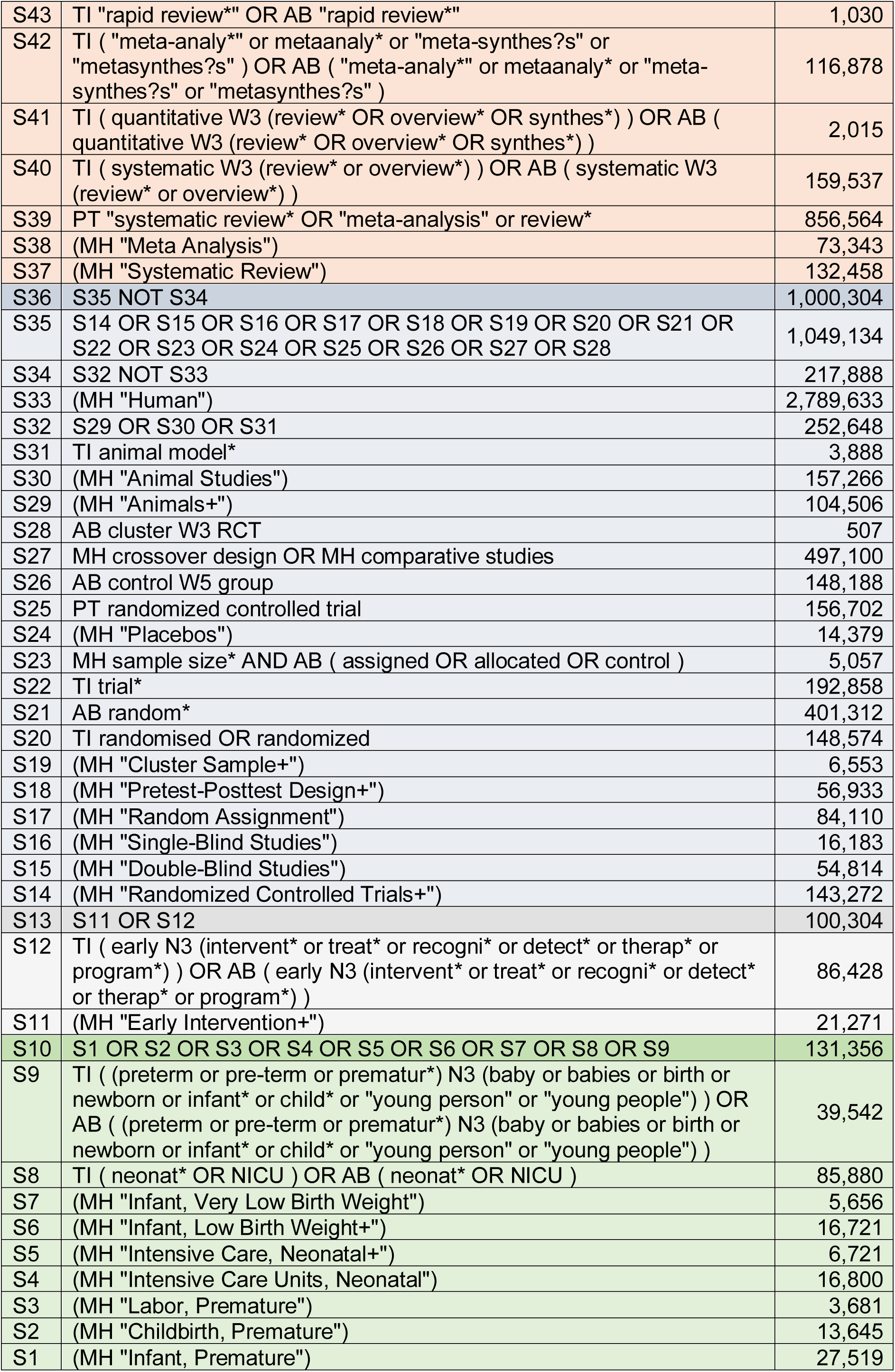
CINAHL Plus with Full Text 07/03/2024.

|  |  |  |
| --- | --- | --- |
| S69 | S67 AND S68 | 14 |
| S68 | S10 AND S59 AND S64 | 316 |
| S67 | S65 AND S66 | 1,628 |
| S66 | S36 OR S44 | 1,735,989 |
| S65 | S10 AND S13 | 3,705 |
| S64 | S60 OR S61 OR S62 OR S63 | 214,303 |
| S63 | TI ( (integrated or cooperat* or co-operat* or coordinat* or co-ordinat* or collaborat* or continuity) N3 (care or therap* or restor* or organi?ation or staff* or approach* or communic* or program* or practice* or manag* or service* or system*) ) OR AB ( (integrated or cooperat* or co-operat* or coordinat* or co-ordinat* or collaborat* or continuity) N3 (care or therap* or restor* or organi?ation or staff* or approach* or communic* or program* or practice* or manag* or service* or system*) ) | 90,964 |
| S62 | TI ( multidisciplinary or "multi-disciplinary" or multiprofessional* or "multi-professional*" or interdisciplinary or "inter-disciplinary" or interprofession* or transdisciplinary or "trans-disciplinary" or transprofessional or "trans-professional" or multimodal or "multi-modal" or multiprofessional or "multi-professional" ) OR AB ( multidisciplinary or "multi-disciplinary" or multiprofessional* or "multi-professional*" or interdisciplinary or "inter-disciplinary" or interprofession* or transdisciplinary or "trans-disciplinary" or transprofessional or "trans-professional" or multimodal or "multi-modal" or multiprofessional or "multi-professional" ) | 97,197 |
| S61 | (MH "Health Care Delivery, Integrated") | 15,410 |
| S60 | (MH "Multidisciplinary Care Team+") | 52,983 |
| S59 | S45 OR S46 OR S47 OR S48 OR S49 OR S50 OR S51 OR S52 OR S53 OR S54 OR S55 OR S56 OR S57 OR S58 | 488,209 |
| S58 | TI speech W1 language OR AB speech W1 language | 10,634 |
| S57 | TI ( "allied health profession*" or AHP* or physiotherap* or "physical therap*" or "occupational therap*" or dietician* or dietetics or psycholog* or "p#ediatric rehabilitation" or therapist* ) OR AB ( "allied health profession*" or AHP* or physiotherap* or "physical therap*" or "occupational therap*" or dietician* or dietetics or psycholog* or "p#ediatric rehabilitation" or therapist* ) | 250,555 |
| S56 | (MH "Rehabilitation, Pediatric") | 1,535 |
| S55 | (MH "Psychologists") | 5,499 |
| S54 | (MH "Psychology+") | 29,623 |
| S53 | (MH "Dietitians") | 6,041 |
| S52 | (MH "Diet Therapy+") | 36,850 |
| S51 | (MH "Rehabilitation, Speech and Language+") | 10,165 |
| S50 | (MH "Speech-Language Pathologists") | 8,179 |
| S49 | (MH "Speech Therapy+") | 4,701 |
| S48 | (MH "Occupational Therapists+") | 10,106 |
| S47 | (MH "Occupational Therapy+") | 27,986 |
| S46 | (MH "Physical Therapists+") | 14,748 |
| S45 | (MH "Physical Therapy+") | 163,150 |
| S44 | S37 OR S38 OR S39 OR S40 OR S41 OR S42 OR S43 | 890,633 |
| S43 | TI "rapid review*" OR AB "rapid review*" | 1,030 |
| S42 | TI ( "meta-analy*" or metaanaly* or "meta-synthes?s" or "metasynthes?s" ) OR AB ( "meta-analy*" or metaanaly* or "meta-synthes?s" or "metasynthes?s" ) | 116,878 |
| S41 | TI ( quantitative W3 (review* OR overview* OR synthes*) ) OR AB ( quantitative W3 (review* OR overview* OR synthes*) ) | 2,015 |
| S40 | TI ( systematic W3 (review* or overview*) ) OR AB ( systematic W3 (review* or overview*) ) | 159,537 |
| S39 | PT "systematic review* OR "meta-analysis" or review* | 856,564 |
| S38 | (MH "Meta Analysis") | 73,343 |
| S37 | (MH "Systematic Review") | 132,458 |
| S36 | S35 NOT S34 | 1,000,304 |
| S35 | S14 OR S15 OR S16 OR S17 OR S18 OR S19 OR S20 OR S21 OR S22 OR S23 OR S24 OR S25 OR S26 OR S27 OR S28 | 1,049,134 |
| S34 | S32 NOT S33 | 217,888 |
| S33 | (MH "Human") | 2,789,633 |
| S32 | S29 OR S30 OR S31 | 252,648 |
| S31 | TI animal model* | 3,888 |
| S30 | (MH "Animal Studies") | 157,266 |
| S29 | (MH "Animals+") | 104,506 |
| S28 | AB cluster W3 RCT | 507 |
| S27 | MH crossover design OR MH comparative studies | 497,100 |
| S26 | AB control W5 group | 148,188 |
| S25 | PT randomized controlled trial | 156,702 |
| S24 | (MH "Placebos") | 14,379 |
| S23 | MH sample size* AND AB ( assigned OR allocated OR control ) | 5,057 |
| S22 | TI trial* | 192,858 |
| S21 | AB random* | 401,312 |
| S20 | TI randomised OR randomized | 148,574 |
| S19 | (MH "Cluster Sample+") | 6,553 |
| S18 | (MH "Pretest-Posttest Design+") | 56,933 |
| S17 | (MH "Random Assignment") | 84,110 |
| S16 | (MH "Single-Blind Studies") | 16,183 |
| S15 | (MH "Double-Blind Studies") | 54,814 |
| S14 | (MH "Randomized Controlled Trials+") | 143,272 |
| S13 | S11 OR S12 | 100,304 |
| S12 | TI ( early N3 (intervent* or treat* or recogni* or detect* or therap* or program*) ) OR AB ( early N3 (intervent* or treat* or recogni* or detect* or therap* or program*) ) | 86,428 |
| S11 | (MH "Early Intervention+") | 21,271 |
| S10 | S1 OR S2 OR S3 OR S4 OR S5 OR S6 OR S7 OR S8 OR S9 | 131,356 |
| S9 | TI ( (preterm or pre-term or prematur*) N3 (baby or babies or birth or newborn or infant* or child* or "young person" or "young people") ) OR AB ( (preterm or pre-term or prematur*) N3 (baby or babies or birth or newborn or infant* or child* or "young person" or "young people") ) | 39,542 |
| S8 | TI ( neonat* OR NICU ) OR AB ( neonat* OR NICU ) | 85,880 |
| S7 | (MH "Infant, Very Low Birth Weight") | 5,656 |
| S6 | (MH "Infant, Low Birth Weight+") | 16,721 |
| S5 | (MH "Intensive Care, Neonatal+") | 6,721 |
| S4 | (MH "Intensive Care Units, Neonatal") | 16,800 |
| S3 | (MH "Labor, Premature") | 3,681 |
| S2 | (MH "Childbirth, Premature") | 13,645 |
| S1 | (MH "Infant, Premature") | 27,519 |

**Table 21.** Cochrane.

|  |  |  |
| --- | --- | --- |
| 1 | MeSH descriptor: [Infant, Premature] explode all trees | 5789 |
| 2 | MeSH descriptor: [Obstetric Labor, Premature] explode all trees | 3305 |
| 3 | MeSH descriptor: [Intensive Care, Neonatal] explode all trees | 428 |
| 4 | MeSH descriptor: [Infant, Low Birth Weight] explode all trees | 3025 |
| 5 | (neonat* or NICU):ti,ab,kw | 29449 |
| 6 | (preterm or pre-term or prematur*) NEAR/3 (baby or babies or birth or newborn or infant* or child* or "young person" or "young people"):ti,ab,kw<br>(Word variations have been searched) | 18880 |
| 7 | OR 1-6 | 40165 |
| 8 | MeSH descriptor: [Early Intervention, Educational] explode all trees | 699 |
| 9 | (early NEAR/3 (intervent* or treat* or recogni* or detect* or therap* or program*):ti,ab,kw<br>(Word variations have been searched) | 35696 |
| 10 | 8 OR 9 | 35696 |
| 11 | 7 AND 10 | 1447 |
| 12 | MeSH descriptor: [Physical Therapists] explode all trees | 261 |
| 13 | MeSH descriptor: [Physical Therapy Modalities] explode all trees | 39290 |
| 14 | MeSH descriptor: [Occupational Therapists] explode all trees | 21 |
| 15 | MeSH descriptor: [Occupational Therapy] explode all trees | 1013 |
| 16 | MeSH descriptor: [Speech Therapy] explode all trees | 417 |
| 17 | MeSH descriptor: [Speech-Language Pathology] explode all trees | 127 |
| 18 | MeSH descriptor: [Nutritionists] explode all trees | 82 |
| 19 | MeSH descriptor: [Diet Therapy] explode all trees | 8203 |
| 20 | MeSH descriptor: [Psychology] explode all trees | 1452 |
| 21 | ("allied health profession" or "allied health professions" or "allied health professional" or "allied health professionals" or AHP* or physiotherap* or "physical therapy" or "physical therapist" or "physical therapists" or "occupational therapy" or "occupational therapist" or "occupational therapists" or dietician* or dietetics or psycholog* or "pediatric rehabilitation" or "paediatric rehabilitation" or therapist*):ti,ab,kw<br>(Word variations have been searched) | 177,105 |
| 22 | (speech NEAR/1 language):ti,ab,kw<br>(Word variations have been searched) | 7575 |
| 23 | MeSH descriptor: [Allied Health Personnel] explode all trees | 1755 |
| 24 | OR 12-23 | 212,004 |
| 25 | MeSH descriptor: [Patient Care Team] explode all trees | 2292 |
| 26 | MeSH descriptor: [Delivery of Health Care, Integrated] explode all trees | 589 |
| 27 | ((multidisciplinary or multi-disciplinary or multiprofessional* or multi-professional* or interdisciplinary or inter-disciplinary or interprofession* or transdisciplinary or trans-disciplinary or transprofessional or trans-professional or multimodal or multi-modal or multiprofessional or multi-professional)):ti,ab,kw<br>(Word variations have been searched) | 23,221 |
| 28 | ((integrated or cooperat* or co-operat* or coordinat* or co-ordinat* or collaborat* or continuity) NEAR/3 (care or therap* or restor* or organi?ation or staff* or approach* or communic* or program* or practice* or manag* or service* or system*)):ti,ab,kw<br>(Word variations have been searched) | 171031 |
| 29 | OR 25-28 | 53,217 |
| 30 | 7 AND 24 AND 29 | 188 |
| 31 | 11 AND 30 | 31 |

## 8.3 APPENDIX 3: Websites searched for neonatal audits

1. Audit Commission (up to 2015)
2. Audit Wales
3. Audit Scotland
4. National Audit Office
5. Guidelines and Audit Implementation Network N.I. (up to 2015).
6. Regulation and Quality Improvement Authority (N.I.)
7. https://digital.nhs.uk/data-and-information/clinical-audits-and-registries
8. https://www.england.nhs.uk/clinaudit/
9. https://www.hqip.org.uk/a-z-of-nca/
10. https://www.hdruk.ac.uk/access-to-health-data/health-data-research-innovation-gateway/

## Footnotes

* This section has been completed by the Centre for Health Economics & Medicines Evaluation (CHEME), Bangor University

